# Disentangling non-linear and time-varying effects in assessing the short-term impact of air pollution on mortality: evidence from a 12-year study in a high-risk Italian area

**DOI:** 10.1101/2025.07.11.25331375

**Authors:** Chiara Marzi, Daniela Nuvolone, Michela Baccini

**Author notes:** Corresponding author: Michela Baccini, Viale Morgani 59, 50134, Florence (FI), Italy. E-mail addresses Chiara Marzi; Daniela Nuvolone; Michela Baccini.

## Abstract

**Introduction:** current evidence on the short-term effects of air pollution on mortality often overlooks potential temporal variation and non-linear exposure–response relationships, which may bias effect estimates and limit the accuracy of health risk assessments.

**Methods:** this study addresses these gaps by examining temporal changes and non-linear associations between daily concentrations of PM_10_, PM_2.5_, NO_2_, and SO_2_ and mortality from natural, cardiovascular, and respiratory causes across eight municipalities in Tuscany, Italy, from 2008 to 2019. Environmental and mortality data were obtained from official sources; missing environmental data were handled through multiple imputation. Time-invariant and time-varying linear effects were estimated using Poisson regression models, and non-linear dose–response curves were assessed using splines.

**Results:** PM_2.5_ and SO_2_ were positively associated with natural and respiratory mortality, while PM_10_ and NO_2_ showed weaker or no associations. Stronger effects were observed during 2012–2015, despite lower pollutant concentrations. SO_2_ also exhibited a non-linear relationship with cardiovascular mortality, with greater effects at lower concentrations.

**Conclusion:** these findings suggest that reductions in pollutant levels do not necessarily imply reduced health risks, potentially due to changes in pollutant composition or interactions with meteorological factors. This study underscores the importance of accounting for both temporal variation and potential non-linearity in air pollution health impact assessments.

**What is already known on this topic:** There is strong evidence that short-term exposure to air pollutants such as PM_10_, PM_2.5_, NO_2_, and SO_2_ is associated with increased risks of natural, cardiovascular, and respiratory mortality. However, most existing studies rely on the assumption that these effects are constant over time and that the exposure–response relationship is linear. The possibility that health effects vary across time due to changing environmental or contextual factors - and that such variation may be non-linear - has received limited attention.

**What this study adds:** This study investigates short-term mortality effects of air pollution over a 12-year period in eight municipalities in Tuscany, Italy, using both linear and non-linear models. It demonstrates that the effects of PM_10_, PM_2.5_, and SO_2_ on mortality are not temporally constant and that stronger associations are observed in periods with lower average pollutant levels. While SO_2_ shows a clear non-linear relationship, non-linearity alone does not fully explain the time variation observed for particulates, suggesting that changes in pollutant composition or environmental conditions may also play a role.

**How this study might affect research, practice or policy:** By revealing that pollutant-related health risks can vary significantly over time and may not follow a simple linear pattern, this study underscores the importance of integrating temporal variability and non-linearity into air pollution epidemiology. These insights could improve the accuracy of health impact assessments, support more responsive air quality regulations, and inform future policies aimed at protecting public health under evolving environmental conditions.

## 1 Introduction

Air pollution is a major public health concern linked to adverse health outcomes and increased mortality rates (1). Over the past decades, many positive and effective measures have been taken to reduce the concentration of air pollutants. However, even at low concentrations, pollutants can still impair human health (2).

Understanding the short-term relationship between air pollution and adverse health outcomes – particularly mortality - is crucial for assessing the immediate health effects of exposure to particulate matter (PM) and other pollutants. A vast body of epidemiological research has consistently linked short-term exposure to outdoor air pollutants - such as PM_10_ and PM_2.5_ (particulate matter with diameters of 10 and 2.5 micrometers or less, respectively), nitrogen dioxide (NO_2_), and sulfur dioxide (SO_2_) - with both total and cause-specific mortality (3–16).

In this context, understanding how the association between air pollution and mortality evolves over time is essential, particularly given potential shifts in pollutant composition and/or contextual factors that may influence effect sizes over long-term observational windows. However, despite the importance of this kind of analysis, relatively few studies have examined the variability of short-term air pollution health effects over extended periods (17–23), often yielding contrasting results. Additionally, from a methodological perspective, a time-varying short-term effect of air pollution on health outcomes could be related also to a violation of the strong but usual assumption that the exposure-response relationship is log-linear (24–28). This assumption implies that any increase in pollutant levels leads to a proportional percentage increase in health events, regardless of the initial exposure level, but whether the relationship remains strictly log-linear across different pollutant concentration ranges is an open question. As a consequence, we cannot exclude that, if the exposure level is not constant across the years, variations of the effect over time could arise from violations of the log-linearity of the exposure-response function rather than from actual effect modifications related to contextual conditions.

In light of this, simultaneously investigating potential non-linearities of the dose-response function and temporal variations of the effect could provide deeper insights into the complexities of air pollution’s impact on public health and enhance the effectiveness of intervention strategies.

This study aims to investigate the short-term effects of PM_10_, PM_2.5_, NO_2_, and SO_2_ on mortality from all natural, cardiovascular, and respiratory causes in the area surrounding Florence (Italy), including eight municipalities, for a total of 597740 inhabitants in 2019, over a period of 12 years, from 2008 to 2019. The area surrounding Florence is a key region in Tuscany for air quality studies due to its distinctive environmental and demographic characteristics. This area is densely populated and highly urbanized, featuring significant industrial activities, heavy traffic, and other sources of pollution that contribute to high concentrations of air pollutants. Moreover, the region’s geographical features, such as valleys and limited air circulation in certain zones, exacerbate pollution levels, leading to the persistent accumulation of harmful particles. These factors make the area particularly vulnerable to the impacts of air pollution. Previous studies in Tuscany have focused on the long-term effects of air pollution (29–31). Recently, a regional project (https://www.acab-toscana.it) quantified the long-term impacts of air pollution on cancer incidence in terms of attributable deaths and years of life lost, but without producing local estimates of exposure-response relationships (32). Regarding short-terms effects, the cities of Florence and Pisa have been included in national meta-analyses (33–39), but no studies to date have assessed these effects over an extended time period, nor have they explored the temporal variability of the associations.

In this study, we investigated the short-term effects of air pollutants on mortality by employing both linear and non-linear dose-response models, with a specific focus on the temporal variation of these associations. Prior to the analysis, we carefully constructed the historical exposure time series, incorporating multiple imputation to address missing data and evaluating the temporal consistency and comparability of pollutant measurements.

## 2 Methods

### 2.1 Data

Daily time series data on air pollution and mortality were collected from 2008 to 2019 for eight municipalities in Tuscany: Bagno a Ripoli, Calenzano, Campi Bisenzio, Florence, Lastra a Signa, Scandicci, Sesto Fiorentino, and Signa (Fig. 1). In this study, we named this area as “Piana Fiorentina”.

**Figure 1.**
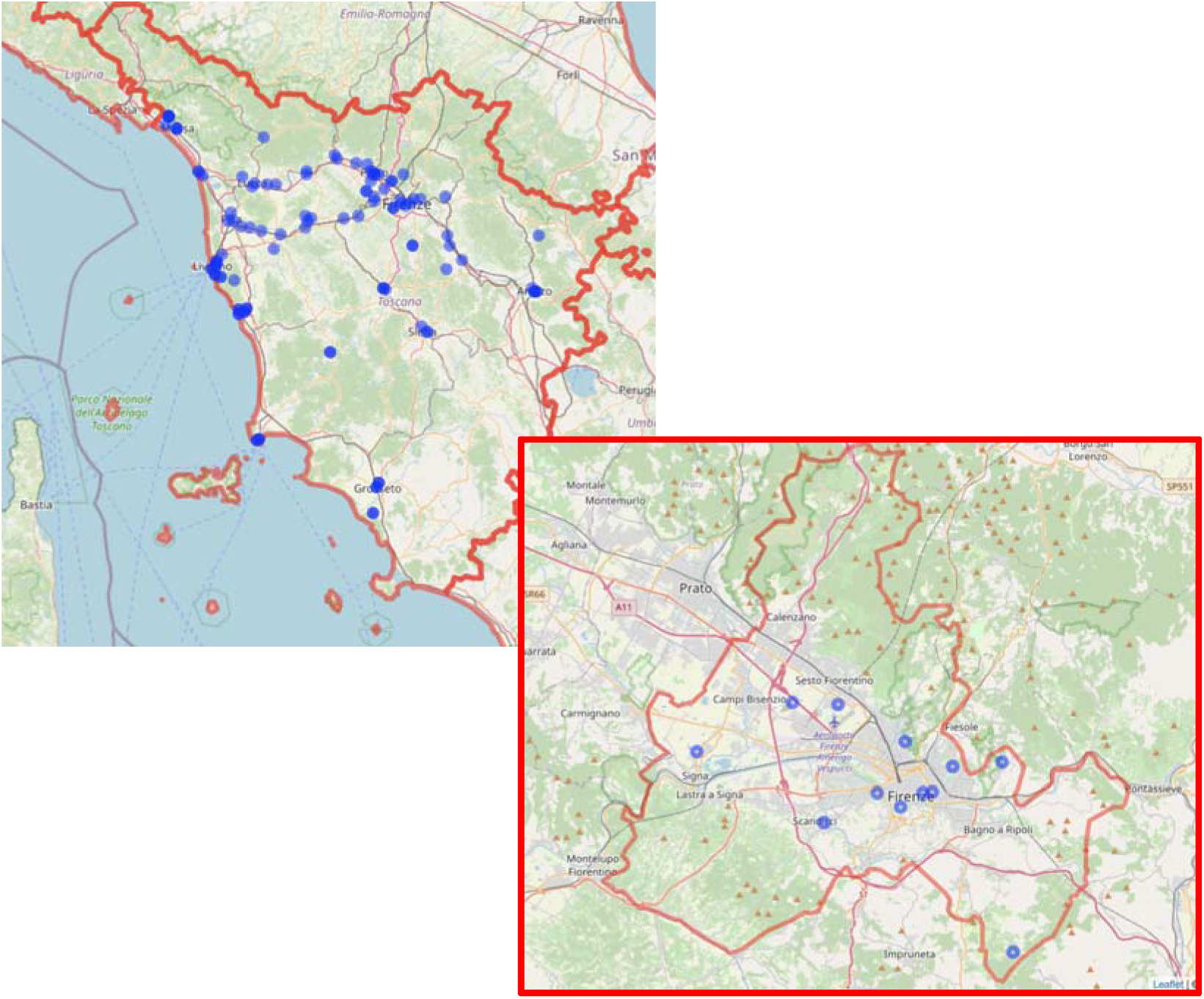
Geographical distribution of air quality monitoring stations across the Tuscany region and the “Piana Fiorentina” area (zoomed-in section). Municipal boundaries are defined according to ISTAT 2019 data.

Mortality data, sourced from mortality registers of the Regional Health Agency of Tuscany (ARS Toscana, https://www.ars.toscana.it), encompassed all natural causes, excluding the external ones (International Classification of Diseases, Ninth Revision (ICD-9), codes below 800, and ICD-10 (Tenth Revision) codes not prefixed with “S” or “T”). We further categorized mortality into cardiovascular (ICD-9 codes 390–459, and ICD-10 codes I00–I99) and respiratory (ICD-9 codes 460–519, and ICD-10 codes J00–J99) causes. We focused on the daily number of deaths among residents of the aforementioned areas, regardless of where the deaths occurred.

Air pollution and, more in general, environmental data used in this study come from a larger dataset obtained collecting data from the monitors belonging to the air quality monitoring network of the Tuscany Regional Environmental Protection Agency (ARPA), meteorological data, including temperature and relative humidity, from the Tuscany Regional Hydrological Service (SIR) and from the measurement sites of the Laboratory of Monitoring and Environmental Modelling for Sustainable Development (LaMMA Consortium). The dataset covers the entire Tuscany region, includes air pollutants beyond PM_10_, PM_2.5_, NO_2_, and SO_2_, and covers a time window larger than the study period (from 2008 to 2019) (Supplementary Table S1). We focused on daily values, computing daily summaries when sub-daily measurements were available (see Supplementary section “Environmental data preprocessing” for further details). Leveraging the air quality data collected by the comprehensive network of sensors across the Tuscany region, we successfully reconstructed the missing PM_10_, PM_2.5_, NO_2_, and SO_2_ data for the days where measurements were unavailable (details provided in section 2.2.1 *Multiple imputation of missing values*).

This study did not involve patients or the local community in its design, conduct, or reporting. However, we intend to engage local communities and policy stakeholders in the dissemination of the findings.

### 2.2 Statistical Analyses

#### 2.2.1 Multiple imputation of missing values

The air quality data collected in Tuscany from 2008 to 2022 included approximately 1,475 variables, each with less than 85% missing values. Overall, 46% of the data were missing, primarily due to long-term station inactivity. To manage computational load, the data were split into five three-year periods (2008–2010, 2011–2013, etc.). Within each period, variables with a missing percentage ≥ 99% were removed, resulting in reduced missingness levels (6.3%, 18%, 18.2%, 10.2%, 3% for each period, respectively). We performed multiple imputation by chained equations (MICE) independently on each three-year period, under a missingness-at-random (MAR) assumption (40). We used CART models to impute missing values, allowing for nonlinearity and interaction effects (41–43). In each CART model, we included as predictors only the variables with less than 35% missing values and a correlation with the specific outcome to be imputed that exceeded a predefined threshold. For air pollution outcomes, we set the minimum correlation between the outcome and predictors to 0.1, following the default of the R *quickpred* function. In contrast, predictors for relative humidity were required to have a correlation of at least 0.7 with the outcome, and those for temperature needed a correlation of at least 0.95. This approach ensured that the CART models contained sufficient number of relevant predictors to maintain imputation accuracy, while avoiding overly complex models that would substantially increase computational time (42). We set the number of imputed datasets to 5, and, within each imputation, we performed 20 iterations to ensure algorithm convergence (44).

After imputing the missing data using all available information from the Tuscany region, we merged environmental and health data. Since mortality data were only available up to December 31, 2019, we focused exclusively on the time period from 2008 to 2019. We selected meteorological stations that measured PM_10_, PM_2.5_, NO_2_, SO_2_, and O3, temperature, and relative humidity within the study area (details about the number and types of monitoring stations were reported in supplementary table S2). For PM_2.5_, both monitoring stations located within the study area were inactive prior to January 1, 2010; therefore, analyses involving PM_2.5_ were limited to the 2010–2019 period. We then aggregated the data to obtain for each air pollutant unique daily time series of average concentrations in the area of interest, along with unique time series of daily average values of temperature and relative humidity. Each imputed dataset underwent statistical analyses separately, and we pooled the five results using a multivariate version of Rubin’s Rule (44) (details in supplementary section “Pooling effect estimates and standard errors”).

#### 2.2.2 Linear effect estimates

Separately for each air pollutant (*z*), we specified Poisson regression models to estimate its short-term effect on daily mortality counts (*Y_t_*), adjusting for confounders. In a first analysis, we assumed a linear effect on a logarithmic scale, that remained constant throughout the study period. To estimate this time-invariant effect we included a linear term for the pollutant concentration averaged over the current and previous day (lag 0–1). We introduced a natural cubic regression spline on calendar day with 5 degrees of freedom per year (*s*(*t*,60)) to account for seasonality and long-term time trend. We adjusted for day of the week and holiday, by including in the model appropriate dummy variables. To control for meteorological conditions, we defined a natural cubic regression spline with 5 degrees of freedom on the average temperature in the current and previous three days (lag 0–3, *s*(*temp_t_*,5)), and included in the model linear and quadratic terms for relative humidity (lag 0-3) (45). The resulting model is the following (Eqs. 1 and 2):

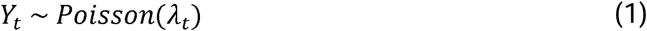

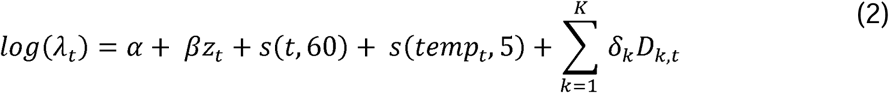

where beta and delta are unknown regression coefficients and the terms D_k,t_ generically indicate the confounders included in the linear predictor.

Successively, given the availability of 12 years of daily measurements, we investigated if and how the effect of the air pollutants on mortality changed over time, maintaining the linear assumption. Time-varying effects were evaluated using two different linear models of increasing complexity: the first model included an interaction term between pollutant concentration and the four-year period (2008–2011 as the reference, 2012–2015, and 2016–2019). In the second model, we introduced a time-varying coefficient for the pollutant (*β_t_*) (Eq. 3):

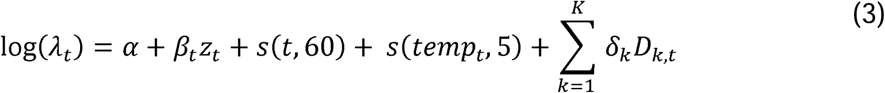

*β_t_* was modeled as combination of a linear coefficient (a) and a natural regression spline on calendar days with 6 degrees of freedom (Eq. 4):

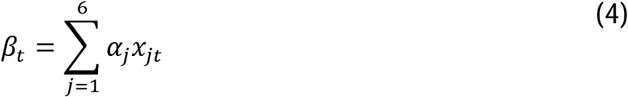

where *x_jt_* represents the *j^th^* basis function of a natural cubic regression spline on calendar time, modelled with 6 degrees of freedom (*s*(*t*,6)). We estimated coefficients *α_j_*, which were then linearly combined with the basis functions of the spline on time to obtain the time-varying coefficient *β_t_* for the pollutant. After estimating both time-invariant and time-varying linear effects in each imputed dataset, the final point estimates of the pollutant effects were obtained by averaging the coefficients across the five imputations. The 90% confidence intervals (CI) have been obtained by accounting for both within-imputation and between-imputation variances, according to the Rubin’s rule (details in supplementary section “Pooling effect estimates and standard errors”). We expressed the effect estimates as percent variations in the number of deaths associated with an increase in pollutant concentration equal to the interquartile range computed in the last year of observation, i.e. 2019.

#### 2.2.3 Non-linear effect estimates

Non-linear effects were assessed by replacing the linear term for the pollutant in Eq. 1 with a natural cubic regression spline with five internal knots at the sextiles of pollutant concentrations (Eq. 5).

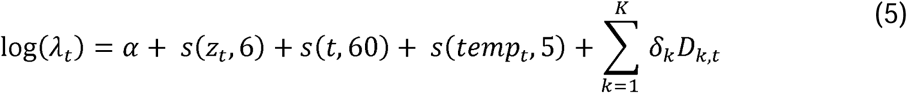

The non-linear relationship between air pollutant and mortality was reported on logarithm scale along with its pointwise 90% confidence bands. We performed this analysis on the whole period and separately for the time windows 2008-2011, 2012-2015, 2016-2019.

#### 2.2.4 Sensitivity analyses

In the linear models, we first investigated the impact of different lag structures for the pollutant exposure by examining a delayed effect (lag2-5) and a prolonged effect (lag0-5). Second, we investigated effect modification by age by incorporating an interaction term between pollutant levels and age groups (< 65 years vs. ≥ 65 years). Third, we evaluated sensitivity to confounding adjustment by modifying the functional form adopted in the model for temperature, relative humidity, and seasonality.

In the non-linear models, we examined the sensitivity of the dose-response relationship by specifying for the air pollutant a spline with three internal knots positioned at the quartiles of the pollutant distribution. This approach allowed us to assess whether the estimated non-linear association remained stable under a more constrained functional form.

Finally, we repeated all analyses excluding monitoring stations that exhibited prolonged gaps in data, specifically those with missing values spanning one or more consecutive years (supplementary Table S2). This sensitivity analysis aimed to ensure that the estimated average pollutant concentrations in the study area - and the resulting exposure metrics - were not biased by stations with extended periods of inactivity clustered in time.

#### 2.2.5 Computational environment

All analyses were conducted on a MacBook Air with an Apple M1 (2020) processor, using R version 4.3.1 within RStudio. The primary statistical analyses and visualizations were performed using the following R packages: mice (v3.16.0) for multiple imputation, mgcv (v1.8.42) for generalized additive modeling, and ggplot2 (v3.5.1) for data visualization.

## 3 Results

Table 1 summarizes daily average atmospheric pollutants concentrations and daily average temperature and humidity from 2008 to 2019 in the study area. Daily concentrations of PM_10_ vary from 2 to 115 *µg/m*^3^, with an average of 27.16 *µg/m*^3^, while daily concentrations of PM_2.5_ vary from 3 to 78.5 *µg/m*^3^, with an average of 16.0 *µg/m*^3^. The daily concentrations of NO_2_ range from 8.9 to 120.8 *µg/m*^3^, with an average of 44.6 *µg/m*^3^, while the daily concentrations of SO_2_ range from values very close to zero to 13.2 *µg/m*^3^, with an average of 1.7 *µg/m*^3^. During the study period, the recommended 24-hour average thresholds defined in the 2021 WHO guidelines have been exceeded on average 36.8, 142.0, 320.7, and 0 days per year for PM_10_, PM_2.5_, NO_2_, and SO_2_, respectively (details in Supplementary Table S3). The averages of the daily mean temperatures and relative humidities were 15.6°C and 64.9%, respectively. In the period 2008-2019 the total number of natural deaths were 76960, with 27156 cardiovascular deaths and 7350 respiratory deaths (Table 1). A graphical representation of the time series of pollutants, meteorological conditions, and daily deaths is shown in Supplementary Fig. S3.

**Table 1.** Descriptive summary for daily air pollutants, meteorological factors, and mortality counts in the Piana Fiorentina area, Italy, during 2008–2019.

| Variable | Mean (SD) | Min | Max | Total |
| --- | --- | --- | --- | --- |
| *Air pollutant* |  |  |  |  |
| **PM<sub>10</sub> ($\mu\text{g}/\text{m}^3$)** | 27.2 (14.4) | 2.5 | 115.2 | - |
| 2008-2011 | 33.9 (15.7) | 8.3 | 115.2 | - |
| 2012-2015 | 25.7 (13.0) | 5.8 | 107.0 | - |
| 2016-2019 | 21.9 (11.5) | 2.5 | 109.0 | - |
| **PM<sub>2.5</sub> ($\mu\text{g}/\text{m}^3$)** | 16.0 (9.5) | 3.0 | 78.5 | - |
| 2010-2011 | 18.1 (10.4) | 5.5 | 78.5 | - |
| 2012-2015 | 16.4 (9.7) | 4.0 | 77.0 | - |
| 2016-2019 | 14.5 (8.6) | 3.0 | 78.0 | - |
| **NO<sub>2</sub> ($\mu\text{g}/\text{m}^3$)** | 44.6 (17.3) | 8.9 | 120.8 | - |
| 2008-2011 | 57.4 (14.8) | 18.8 | 120.8 | - |
| 2012-2015 | 42.6 (15.6) | 12.2 | 101.0 | - |
| 2016-2019 | 34.0 (12.6) | 8.9 | 78.4 | - |
| **SO<sub>2</sub> ($\mu\text{g}/\text{m}^3$)** | 1.7 (1.4) | 0.0 | 13.2 | - |
| 2008-2011 | 1.3 (0.7) | 0.2 | 5.4 | - |
| 2012-2015 | 1.9 (1.8) | 0.0 | 13.2 | - |
| 2016-2019 | 1.7 (1.3) | 0.0 | 10.4 | - |
| *Meteorological factor* |  |  |  |  |
| **Temperature (°C)** | 15.8 (7.55) | -4.4 | 34.0 | - |
| 2008-2011 | 15.1 (7.4) | -4.4 | 30.6 | - |
| 2012-2015 | 15.7 (7.2) | -2.7 | 30.8 | - |
| 2016-2019 | 16.4 (8.0) | -1.2 | 34.5 | - |
| **Relative humidity (%)** | 64.9 (15.6) | 20.2 | 100.0 | - |
| 2008-2011 | 66.6 (14.9) | 28.0 | 100.0 | - |
| 2012-2015 | 65.2 (15.6) | 26.2 | 100.0 | - |
| 2016-2019 | 62.8 (16.2) | 20.2 | 99.6 | - |
| *Mortality counts* |  |  |  |  |
| **Natural deaths** | 17.6 (4.9) | 3 | 49 | 76960 |
| 2008-2011 | 17.4 (4.7) | 6 | 39 | 25460 |
| 2012-2015 | 17.5 (5.1) | 5 | 49 | 25521 |
| 2016-2019 | 18.1 (5.0) | 3 | 38 | 25979 |
| **Cardiovascular deaths** | 6.2 (2.7) | 0 | 23 | 27156 |
| 2008-2011 | 6.5 (2.7) | 1 | 18 | 9435 |
| 2012-2015 | 6.2 (2.7) | 0 | 23 | 8996 |
| 2016-2019 | 6.1 (2.7) | 0 | 18 | 8725 |
| **Respiratory deaths** | 1.7 (1.4) | 0 | 10 | 7350 |
| 2008-2011 | 1.6 (1.4) | 0 | 10 | 2342 |
| 2012-2015 | 1.6 (1.4) | 0 | 9 | 2282 |
| 2016-2019 | 1.9 (1.5) | 0 | 9 | 2726 |

Max: maximum, Min: minimum, SD: standard deviation.

### 3.1 Linear effect estimates

A linear and time-invariant effect of PM_2.5_ and SO_2_ on mortality was observed (Table 2). Specifically, a 7.5 *µg/m*^3^ increase in PM_2.5_ was associated with a 0.87% increase in natural mortality (90% CI: 0.17, 1.57), while a 0.98 *µg/m*^3^ increase in SO_2_ concentration corresponded to a 1.65% increase in natural mortality (90% CI: 0.92, 2.38) and a 3.69% increase in respiratory mortality (90% CI: 1.53, 5.90). In contrast, the estimates of the time-invariant linear effect showed no associations between PM_10_ or NO_2_ exposure and natural, cardiovascular, or respiratory mortality. For a 10.5 *µg/m*^3^ increase in PM_10_, the estimated percent changes were 0.46 (90% CI: -0.20, 1.12) for natural mortality, -0.28 (90% CI: -1.30, 0.75) for cardiovascular mortality, and -0.16 (90% CI: -2.12, 1.85) for respiratory mortality. For NO_2_, a 13.2 *µg/m*^3^ increase was associated with estimated percent changes of 0.38 (90% CI: - 0.49, 1.25) for natural mortality, -0.34 (90% CI: -1.79, 1.12) for cardiovascular mortality, and -2.89 (90% CI: -5.52, -0.2) for respiratory mortality.

**Table 2.**
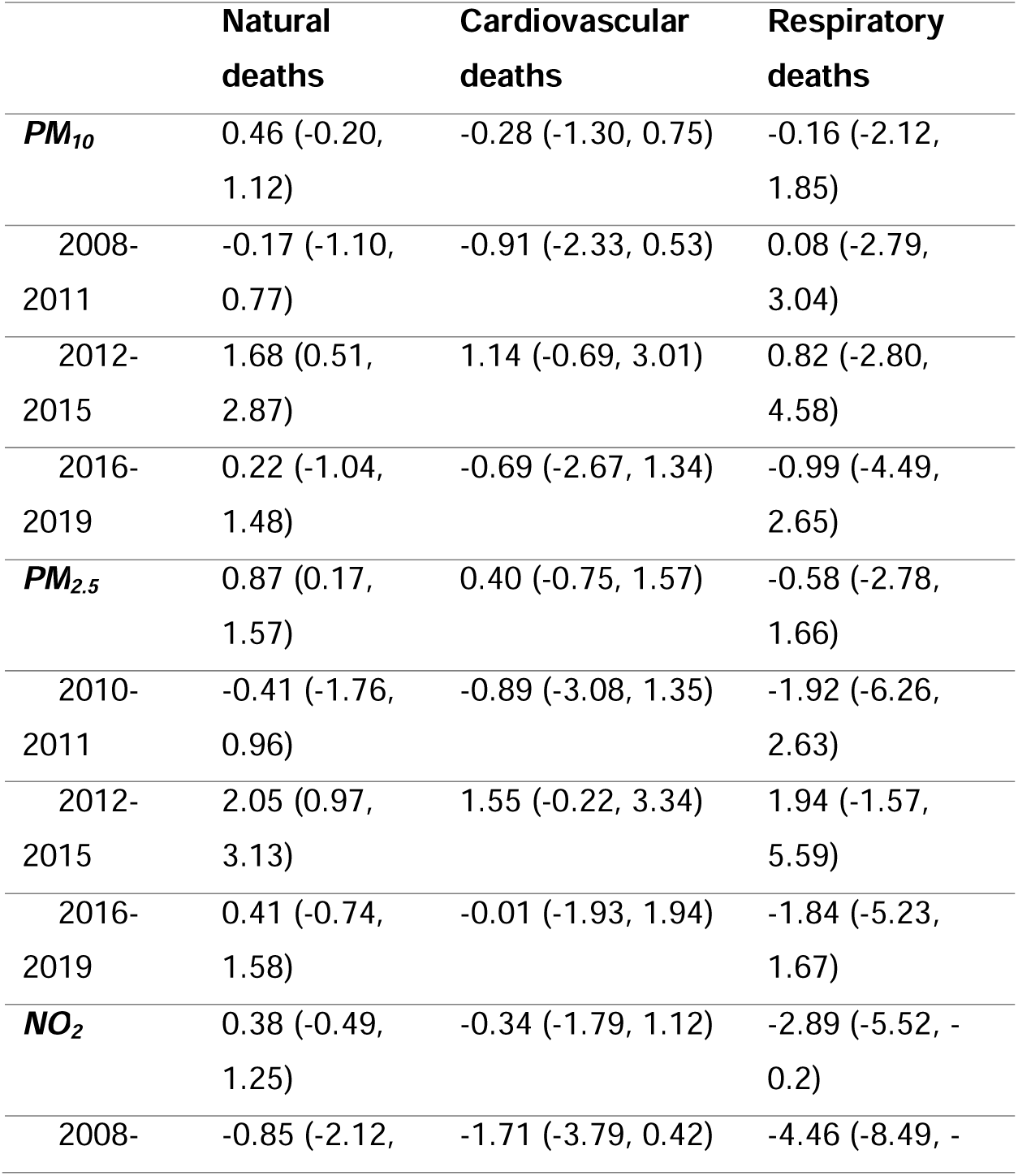
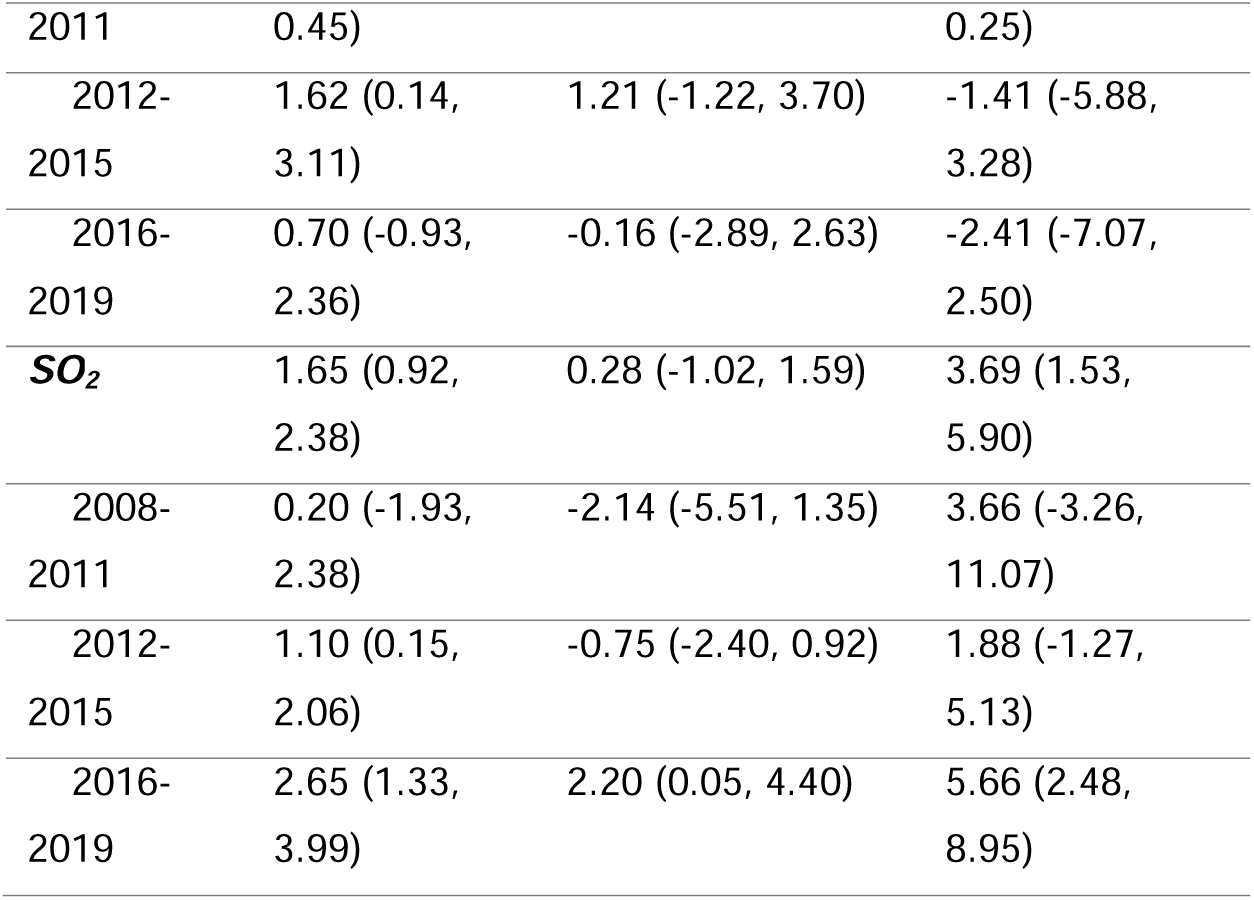
Results from the time-invariant linear model. Estimates and 90% confidence intervals (CI) of the percent variations in the number of deaths associated with an increase in pollutant concentration equal to the **interquartile range computed in 2019 (10.5** *µg⁄m*^3^ **for PM_10_, 7.5** *µg⁄m*^3^ **for PM_2.5_, 13.2** *µg⁄m*^3^ **for NO_2_, 0.98** *µg⁄m*^3^ **for SO_2_).**

The results of the linear models assessing the temporal variability of the effect by the introduction of an interaction term between air pollutant and the 4-years period indicator suggest a change in the impact of PM_10_ and PM_2.5_ on natural mortality during the 2012–2015 period (Fig. 2). Specifically, there is moderate evidence of interaction as indicated by likelihood ratio test (LRT) (p-values<0.1) and the Akaike Information Criterion (AIC) which is lower for the model with interaction compared to models without interaction. During the 2012–2015 period, a 10.5 *µg/m*^3^increase in PM_10_ was associated with a 1.68% increase in natural mortality (90% CI: 0.51, 2.87), while a 7.5 *µg/m*^3^ increase in PM_2.5_ corresponded to a 2.05% increase (90% CI: 0.97, 3.13). Although Figs. 2 and 3 suggest that the effects of NO_2_ on natural mortality also exhibit temporal variability, with a stronger effect observed during the 2012–2015 period, there is insufficient statistical evidence to confirm this pattern (LRT p-values > 0.1 and AIC values higher than those of the linear models without interaction). When assessing the time-varying linear effects using the more flexible model specification, the association between SO_2_ and natural mortality showed a lower AIC value compared to the time-invariant model, indicating a better model fit. This time-varying effect of SO_2_ is also visually evident in Fig. 3j. Regardless of the complexity of the model used to assess temporal variability, for PM_10_, PM_2.5_, and SO_2_, the strongest effects consistently emerged during periods when annual average pollutant concentrations were lower.

**Figure 2.**
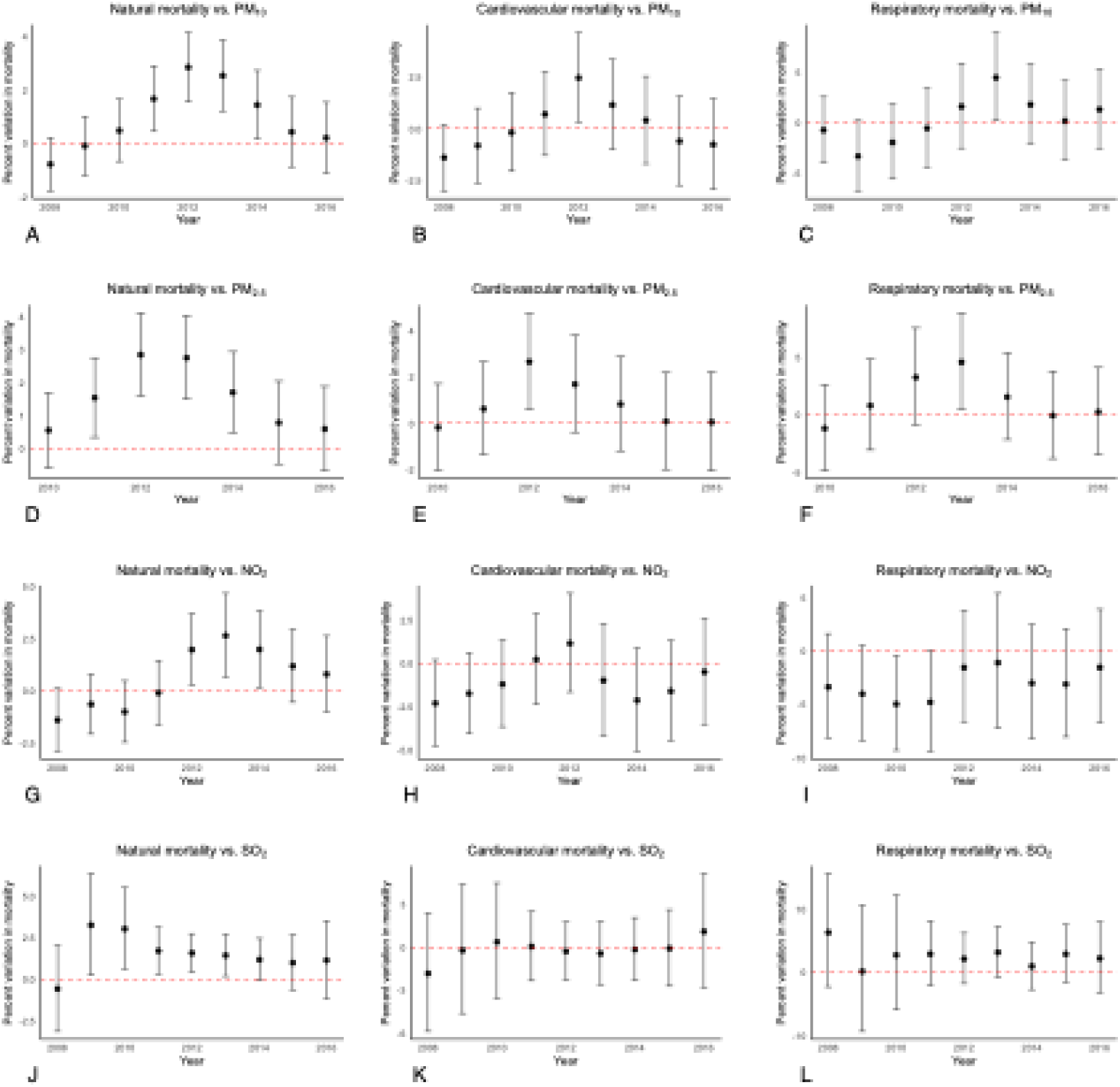
Time-variant linear effects of PM_10_, PM_2.5_, NO_2_, and SO_2_ on mortality. Each dot represents the percent variation in the number of deaths associated with an increase in pollutant concentration equal to the interquartile range computed in the last year of observation (2019), specifically: 10.5 *µg⁄m*^3^ for PM_10_, 7.5 *µg⁄m*^3^ for PM_2.5_, 13.2 *µg⁄m*^3^ for NO₂, and 0.98 *µg⁄m*^3^ for SO₂. The value at each year corresponds to the effect estimated for the four-year period beginning in that year. The whiskers indicate the corresponding 90% confidence intervals. All estimates are pooled over the five imputations.

**Figure 3.**
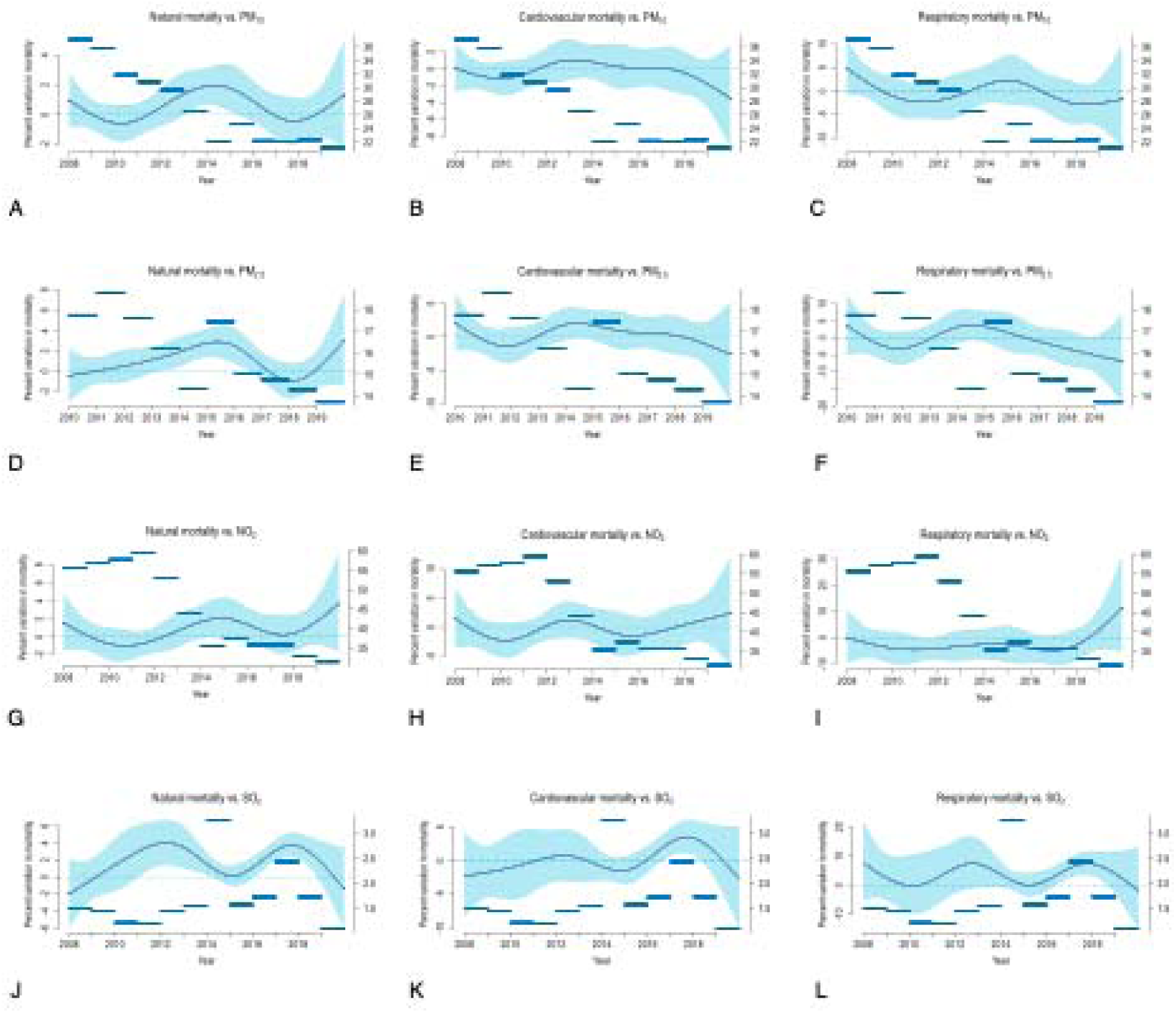
Time-variant linear effects of PM_10_, PM_2.5_, NO_2_, and SO_2_ on mortality. The time-varying coefficient for each pollutant was modelled as the combination of a linear term and a natural regression spline on calendar days with 6 degrees of freedom. Horizontal lines represent the annual average concentration of the corresponding pollutant.

### 3.2 Non-linear effect estimates

In Fig. 4, we graphically illustrate the relationship between air pollution concentration and mortality on a logarithmic scale, modelled using a spline with 6 dof. Although the dose-response curves for natural mortality in relation to PM_10_ and PM_2.5_ exhibit some degree of uncertainty, they suggest slightly stronger effects at lower PM_10_ concentrations (<30 *µg/m*^3^, Fig. 4a) and lower PM_2.5_ concentrations (<20 *µg/m*^3^, Fig. 4d). However, there is no strong evidence of non-linearity, as indicated by LRT p-values > 0.1 and AIC values for the non-linear models exceeding those of the linear models. Similarly, no strong evidence of non-linearity is observed in the relationship between PM_10_ and cardiovascular or respiratory mortality, nor in the association between NO_2_ and all-cause mortality (LRT p-values > 0.1 and AIC values higher for non-linear models than for linear models). In contrast, the relationship between PM_2.5_ and SO_2_ and cardiovascular mortality exhibits clear non-linear patterns, as indicated by LRT p-values < 0.05 and lower AIC values for the non-linear models compared to their linear counterparts. Specifically, the effect of PM_2.5_ follows a parabolic curve, while the association between SO_2_ and cardiovascular mortality is stronger at lower concentration levels (Fig. 4k). However, for SO_2_-related effects on natural and respiratory mortality, there is no substantial evidence of non-linearity, with LRT p-values > 0.1 and AIC values for non-linear models exceeding those of the corresponding linear models.

**Figure 4.**
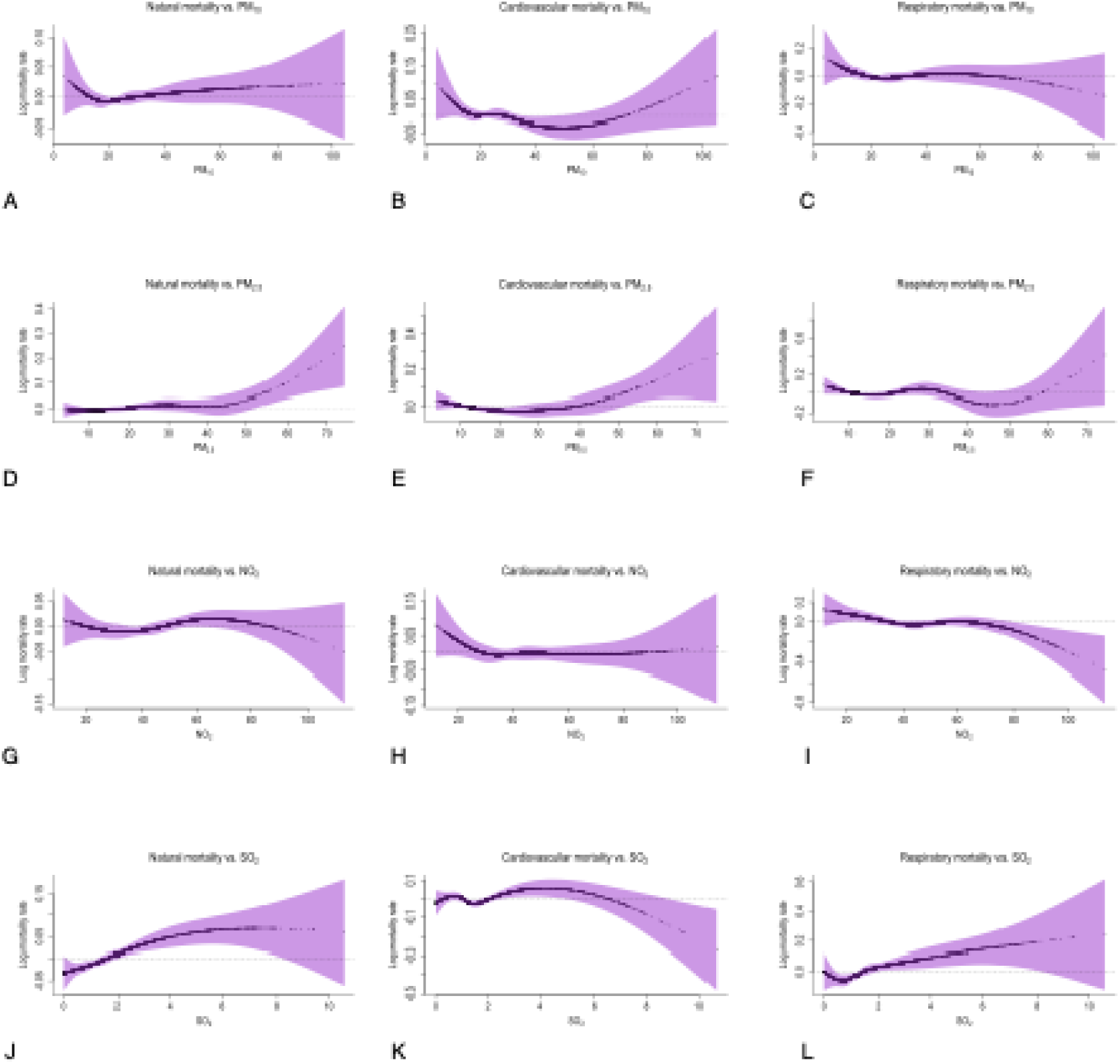
Non-linear effects of PM_10_, PM_2.5_, NO_2_, and SO_2_ on mortality.

When considering the non-linear relationship between natural mortality and particulate or NO_2_ concentration levels across the different four-year periods, it is evident that the trend during 2012–2015 follows a positive trajectory (Fig. 4g, 4h, and 4i), with no evident deviations from linearity (LRT p-values > 0.2). In contrast, for SO_2_, there is some evidence suggesting that its effect on natural mortality may be non-linear during the 2012–2015 period, with stronger effects at lower concentration levels (<2 *µg/m*^3^) (Fig. 4j), as indicated by an LRT p-value ≈ 0.1.

These patterns are consistent with those observed when the analysis is stratified by each time window, i.e., 2008–2011, 2012–2015, and 2016–2019 (Supplementary Figs. S4, S5, S6, and S7).

### 3.3 Sensitivity analyses

The results of the sensitivity analyses confirm the robustness of our findings under different modelling choices and dataset restrictions. In general, the estimated effects remained stable across alternative lag structures, with similar trends observed when considering a delayed effect (lag2-5) or a prolonged effect (lag0-5) (Supplementary Table S4). Similarly, incorporating an interaction term between pollutant levels and age groups (< 65 years vs. ≥ 65 years) did not substantially alter the results, indicating that the overall exposure-response relationship did not differ markedly between age strata (Supplementary Table S5).

Adjustments for confounding variables through modifications in the functional form of temperature, relative humidity, and time did not lead to significant deviations in the estimated effects (Supplementary Table S6). When considering non-linear models, restricting the spline for pollutants to three internal knots positioned at the quartiles of the pollutant distribution resulted in minor variations but did not affect the overall shape of the dose-response curves (Supplementary Fig. S8).

The exclusion of monitoring stations that were inactive for part of the study period yielded consistent results, suggesting that data availability over time did not bias the findings (Supplementary Table S7).

## 4 Discussion

This study assessed the short-term effects of PM_10_, PM_2.5_, NO_2_, and SO_2_ on mortality over a 12-year period. No significant associations were found for PM_10_ or NO_2_ in time-invariant models, whereas PM_2.5_ and SO_2_ showed a consistent association with natural and respiratory mortality (only for SO_2_). Temporal variation in the effects of PM_10_, PM_2.5_, and SO_2_ on natural mortality was observed, with stronger associations at lower concentrations. While no strong evidence of non-linearity was found for PM_10_, PM_2.5_, or NO_2_, SO_2_ exhibited a non-linear relationship with natural mortality during the 2012–2015 period.

Our findings on positive time-invariant associations between daily PM_2.5_ and SO_2_ levels and mortality align with previous studies (7,8,22,46–48). Although the results for PM_10_ and NO_2_ show less marked effects, the estimated associations still follow the general trend described in earlier research, indicating a positive direction of impact (7,8,22,45,46,49–56).

Although we observed a decreasing trend in air pollution concentrations, consistent with trends reported in other regions,(18–23) this improvement - likely due to stricter regulations - health effects did not decrease accordingly. This aligns with studies suggesting do not always reduce health risks and may coincide with increased toxicity, particularly of airborne particles (57).

As previously raised by other authors (19,23,58), the observed time variability in health effects may partly be explained by a non-linear dose-response relationship between pollutant concentrations and mortality. This seems plausible for SO_2_-related mortality effects, as its dose-response function exhibited a deviation from log-linearity, showing steeper effects at lower levels. In contrast, we found no clear evidence of non-linearity for PM_10_ and PM_2.5_ that could alone account for their time-varying effects. Instead, we hypothesize that this variation may be due to changes in particulate composition over time, potentially resulting in increased toxicity.

Unlike other ambient risk factors such as SO_2_, or NO_2_, PM is a heterogeneous mix of chemically, physically, and often biologically diverse components. This heterogeneity means that the same PM mass concentration can have different toxicological properties depending on its composition. Evidence shows that combustion- and traffic-related components - such as black carbon, nitrate, sulfate, calcium, road dust, and metals like cadmium, lead, vanadium, and zinc - are particularly harmful (59–63). Ultrafine particles (UFPs), under 100 nanometers in diameter, may be especially toxic (64), with number concentration (i.e., number of particles per unit volume of air) possibly more relevant than mass concentration for health effects (65–69). Although our study lacked PM composition data, several factors likely influenced PM toxicity in the study area. These include ongoing urbanization and major infrastructure developments, such as the expansion of public transport systems (2011-2017) and the construction of a new high-speed rail line and station (began in 2010), which likely influenced both population density and traffic patterns. Furthermore, in 2010, a local ordinance (Ordinanza 2010/00401) restricted the circulation of public transport vehicles not compliant with at least Euro 2 diesel emission standards, potentially modifying the fleet composition and emissions. During the study period, diesel vehicles in Florence province rose from 44% to 54%, with 23% still below Euro 5 in 2019 (from Automobile Club d’Italia, ACI: https://aci.gov.it/attivita-e-progetti/studi-e-ricerche/autoritratto/). This shift may have increased PM toxicity, counteracting benefits from emission reductions. A similar mechanism was suggested in a Rome-based study (22).

Another important aspect to explore is the potential interaction between particulate exposure and temperature, because temperature may influence the toxicity of particulate matter (PM), particularly through its role in the formation of secondary pollutants such as sulfate and nitrate. These compounds are produced via photochemical reactions, which are enhanced by higher temperatures and solar radiation (70) - conditions expected to increase with climate change. Understanding the interaction between temperature and PM is thus critical for assessing combined environmental health risks. Moreover, mismatched temporal scales - such as multi-day heatwaves versus daily pollutant effects - can complicate interpretation, especially in the context of harvesting - where acute environmental stressors trigger a temporary rise in mortality among frail individuals, followed by a compensatory decline. In such scenarios, pollutants may misleadingly appear to have a protective effect in the immediate aftermath. Investigating these dynamics is essential for clarifying exposure–response relationships and improving public health interventions.

Our study presents several strengths and some limitations. Notably, we relied on high-quality environmental and health data, with pollutant concentrations derived from direct measurements at validated monitoring stations. Unlike model-based or satellite-derived estimates (see, e.g., (71–73)), which can introduce spatial and temporal uncertainty, this approach provides a more accurate representation of local exposure. Furthermore, missing data were handled using robust multiple imputation techniques, with uncertainty appropriately propagated throughout the analytical workflow.

Several limitations should be acknowledged. First, it focuses on a single geographic area - albeit encompassing multiple municipalities - which may limit the generalizability of findings to other regions with different environmental or sociodemographic contexts. Second, the lack of data on particulate matter composition prevents a direct evaluation of whether changes in particle chemistry over time contributed to increased toxicity and health impacts.

## 5 Conclusion

In conclusion, this study highlights the complexity of the short-term effects of air pollution on mortality, showing that while overall concentrations have decreased, health risks have not followed the same downward trend. The observed temporal variations - particularly for particulate matter and SO_2_ - underscore the importance of considering pollutant composition, meteorological interactions, and time-varying toxicity in future assessments. Given that non-linearity and temporal variability may be interrelated and difficult to fully disentangle, future studies should aim to jointly evaluate these aspects to better understand the dynamics of pollutant-related health risks.

## Statements

### Contributorship statement

CM and MB conceptualized the work. CM and DN managed data collection. CM and MB defined the methodology. CM performed formal analysis and visualization. MB acquired funding and supervised the entire work. CM and MB wrote the original draft. All authors review and approved the final manuscript.

During the preparation of this manuscript, the author(s) used OpenAI’s ChatGPT-4o to assist with improving the readability and clarity of the text. All content generated with the help of this tool was subsequently reviewed and edited by the authors, who take full responsibility for the final version of the manuscript.

### Data availability

Data are available upon reasonable request.

### Ethics approval and consent to participate

This study used aggregated mortality data, which contain no personally identifiable information and are publicly available from the Italian National Institute of Statistics.

Therefore, the study does not involve human subjects as defined by ethical review standards, and no ethics committee approval or informed consent was required.

### Consent for publication

Not applicable

## Acknowledgements

The authors gratefully acknowledge the Tuscany Regional Hydrological Service (SIR) and the Laboratory of Monitoring and Environmental Modelling for Sustainable Development (LaMMA Consortium) for providing environmental data essential to this study. Their support and collaboration have been invaluable to the research.

## References

1. WHO Global Air Quality Guidelines: Particulate Matter (PM2. 5 and PM10), Ozone, Nitrogen Dioxide, Sulfur Dioxide and Carbon Monoxide. 1st ed. Geneva: World Health Organization; 2021. 1 p.

2. Di Q, Wang Y, Zanobetti A, Wang Y, Koutrakis P, Choirat C, et al. Air Pollution and Mortality in the Medicare Population. N Engl J Med. 2017 Jun 29;376(26):2513–22.

3. Gyaase S, Nyame S, Klipstein-Grobusch K, Asante KP, Downward GS. Climate, Air Quality and Their Contribution to Cardiovascular Disease Morbidity and Mortality in Low- and Middle-Income Countries: A Systematic Review and Meta-Analysis. gh. 2025 Mar 27;20(1):35.

4. Orellano P, Reynoso J, Quaranta N, Bardach A, Ciapponi A. Short-term exposure to particulate matter (PM10 and PM2.5), nitrogen dioxide (NO2), and ozone (O3) and all-cause and cause-specific mortality: Systematic review and meta-analysis. Environment International. 2020 Sep;142:105876.

5. Luben TJ, Wilkie AA, Krajewski AK, Njie F, Park K, Zelasky S, et al. Short-term exposure to air pollution and infant mortality: A systematic review and meta-analysis. Science of The Total Environment. 2023 Nov;898:165522.

6. Badida P, Krishnamurthy A, Jayaprakash J. Meta analysis of health effects of ambient air pollution exposure in low- and middle-income countries. Environmental Research. 2023 Jan;216:114604.

7. Gariazzo C, Renzi M, Marinaccio A, Michelozzi P, Massari S, Silibello C, et al. Association between short-term exposure to air pollutants and cause-specific daily mortality in Italy. A nationwide analysis. Environ Res. 2023 Jan 1;216(Pt 3):114676.

8. Renzi M, Marchetti S, De’Donato F, Pappagallo M, Scortichini M, Davoli M, et al. Acute effects of particulate matter on all-cause mortality in urban, rural, and suburban areas, Italy. International Journal of Environmental Research and Public Health. 2021;18(24):12895.

9. Mills IC, Atkinson RW, Anderson HR, Maynard RL, Strachan DP. Distinguishing the associations between daily mortality and hospital admissions and nitrogen dioxide from those of particulate matter: a systematic review and meta-analysis. BMJ Open. 2016 Jul;6(7):e010751.

10. Lu F, Xu D, Cheng Y, Dong S, Guo C, Jiang X, et al. Systematic review and meta-analysis of the adverse health effects of ambient PM2.5 and PM10 pollution in the Chinese population. Environmental Research. 2015 Jan;136:196–204.

11. Fajersztajn L, Saldiva P, Pereira LAA, Leite VF, Buehler AM. Short-term effects of fine particulate matter pollution on daily health events in Latin America: a systematic review and meta-analysis. Int J Public Health. 2017 Sep;62(7):729–38.

12. Atkinson RW, Kang S, Anderson HR, Mills IC, Walton HA. Epidemiological time series studies of PM_2.5_ and daily mortality and hospital admissions: a systematic review and meta-analysis. Thorax. 2014 Jul;69(7):660–5.

13. Wang M, Li H, Huang S, Qian Y, Steenland K, Xie Y, et al. Short-term exposure to nitrogen dioxide and mortality: A systematic review and meta-analysis. Environmental Research. 2021 Nov;202:111766.

14. He MZ, Kinney PL, Li T, Chen C, Sun Q, Ban J, et al. Short- and intermediate-term exposure to NO2 and mortality: A multi-county analysis in China. Environmental Pollution. 2020 Jun;261:114165.

15. Requia WJ, Alahmad B, Koutrakis P. Short-term exposure to sulfur dioxide and daily mortality in Brazil: A nationwide time-series study between 2003–2017. Chemosphere. 2023 Dec;343:140259.

16. Chen L, Wang X, Qian Z, Sun L, Qin L, Wang C, et al. Ambient gaseous pollutants and emergency ambulance calls for all-cause and cause-specific diseases in China: a multicity time-series study. Environ Sci Pollut Res. 2022 Apr;29(19):28527–37.

17. Dominici F, Peng RD, Zeger SL, White RH, Samet JM. Particulate Air Pollution and Mortality in the United States: Did the Risks Change from 1987 to 2000? American Journal of Epidemiology. 2007 Aug 28;166(8):880–8.

18. Fischer PH, Marra M, Ameling CB, Janssen N, Cassee FR. Trends in relative risk estimates for the association between air pollution and mortality in The Netherlands, 1992–2006. Environmental Research. 2011 Jan;111(1):94–100.

19. Kim H, Kim H, Lee JT. Effects of ambient air particles on mortality in Seoul: Have the effects changed over time? Environmental Research. 2015 Jul;140:684–90.

20. Nishikawa H, Seposo XT, Madaniyazi L, Kim Y, Tobías A, Yamagami M, et al. Long-term trends in mortality risk associated with short-term exposure to air pollution in 10 Japanese cities between 1977 and 2015. Environmental Research. 2023 Feb;219:115108.

21. Perez L, Grize L, Infanger D, Künzli N, Sommer H, Alt GM, et al. Associations of daily levels of PM10 and NO2 with emergency hospital admissions and mortality in Switzerland: Trends and missed prevention potential over the last decade. Environmental Research. 2015 Jul;140:554–61.

22. Renzi M, Stafoggia M, Faustini A, Cesaroni G, Cattani G, Forastiere F. Analysis of Temporal Variability in the Short-term Effects of Ambient Air Pollutants on Nonaccidental Mortality in Rome, Italy (1998–2014). Environ Health Perspect. 2017 Jun 23;125(6):067019.

23. Schwarz M, Peters A, Stafoggia M, de’Donato F, Sera F, Bell ML, et al. Temporal variations in the short-term effects of ambient air pollution on cardiovascular and respiratory mortality: a pooled analysis of 380 urban areas over a 22-year period. The Lancet Planetary Health. 2024 Sep;8(9):e657–65.

24. Roberts S, Martin MA. The Question of Nonlinearity in the Dose-Response Relation between Particulate Matter Air Pollution and Mortality: Can Akaike’s Information Criterion be Trusted to Take the Right Turn? American Journal of Epidemiology. 2006 Oct 13;164(12):1242–50.

25. Daniels MJ. Estimating Particulate Matter-Mortality Dose-Response Curves and Threshold Levels: An Analysis of Daily Time-Series for the 20 Largest US Cities. American Journal of Epidemiology. 2000 Sep 1;152(5):397–406.

26. Schwartz J, Zanobetti A. Using Meta-Smoothing to Estimate Dose-Response Trends across Multiple Studies, with Application to Air Pollution and Daily Death: Epidemiology. 2000 Nov;11(6):666–72.

27. Daniels MJ, Dominici F, Zeger SL, Samet JM. The National Morbidity, Mortality, and Air Pollution Study. Part III: PM10 concentration-response curves and thresholds for the 20 largest US cities. Res Rep Health Eff Inst. 2004 May;(94 Pt 3):1–21; discussion 23-30.

28. Smith RL, Spitzner D, Kim Y, Fuentes M. Threshold Dependence of Mortality Effects for Fine and Coarse Particles in Phoenix, Arizona. Journal of the Air & Waste Management Association. 2000 Aug;50(8):1367–79.

29. Marabotti C, Piaggi P, Scarsi P, Venturini E, Cecchi R, Pingitore A. Mortality for chronic-degenerative diseases in Tuscany: Ecological study comparing neighboring areas with substantial differences in environmental pollution. Int J Occup Med Environ Health [Internet]. 2017 May 24 [cited 2024 Sep 30]; Available from: http://www.journalssystem.com/ijomeh/Mortality-for-chronic-degenerative-diseases-in-Tuscany-an-ecological-study-comparing-neighbouring-areas-with-substantial-differences-in-environmental-pollution-,63928,0,2.html

30. Minichilli F, Santoro M, Linzalone N, Maurello MT, Sallese D, Bianchi F. Studio epidemiologico di coorte residenziale su mortalità e ricoveri ospedalieri nell’area intorno all’inceneritore di San Zeno, Arezzo. E&P. 2016 Feb;40(1):33–43.

31. Uccelli R, Mastrantonio M, Altavista P, Pacchierotti F, Piersanti A, Ciancarella L. Impact of modelled PM2.5, NO2 and O3 annual air concentrations on some causes of mortality in Tuscany municipalities. European Journal of Public Health. 2019 Oct 1;29(5):871–6.

32. Baccini M, Pirona F, Grisotto L, Cereda G, Lachi A, Levi M, et al. Tracheal, bronchus, and lung cancer mortality and air pollution exposure in Tuscany, Italy: Bayesian Health Impact Assessment and Global Sensitivity Analysis on a sub-regional scale. Environmental Pollution. 2025 Mar;368:125682.

33. Baccini M, Biggeri A, Gruppo collaborativo EpiAir2. [Short-term impact of air pollution among Italian cities covered by the EpiAir2 project]. Epidemiol Prev. 2013;37(4–5):252–62.

34. Baccini M, Biggeri A, Lagazio C, Lerxtundi A, Schwartz J. Comparison of alternative modelling techniques in estimating short-term effect of air pollution with application to the Italian meta-analysis data (MISA Study). Epidemiologia e prevenzione. 2006;30:279–88.

35. Bellini P, Baccini M, Biggeri A, Terracini B. The meta-analysis of the Italian studies on short-term effects of air pollution (MISA): old and new issues on the interpretation of the statistical evidences. Environmetrics. 2007;18(3):219–29.

36. Biggeri A, Baccini M, Bellini P, Terracini B. Meta-analysis of the Italian Studies of Short-term Effects of Air Pollution (MISA), 1990–1999. International Journal of Occupational and Environmental Health. 2005 Jan;11(1):107–22.

37. Biggeri A, Bellini P, Terracini B. [Meta-analysis of the Italian studies on short-term effects of air pollution--MISA 1996-2002]. Epidemiol Prev. 2004;28(4-5 Suppl):4–100.

38. Forastiere F, Faustini A. Short-term effects of air pollution on human health: from epidemiological research to epidemiological surveillance. Epidemiologia e Prevenzione. 2009;33(6 Suppl 1):5–12.

39. Gandini M, Berti G, Cattani G, Faustini A, Scarinzi C, De’donato F, et al. Environmental indicators in EpiAir2 project: air quality data for epidemiological surveillance. Epidemiologia e prevenzione. 2013;37(4–5):209–19.

40. Rubin DB, editor. Multiple Imputation for Nonresponse in Surveys. Hoboken, NJ, USA: John Wiley & Sons, Inc.; 1987. (Wiley Series in Probability and Statistics).

41. Burgette LF, Reiter JP. Multiple imputation for missing data via sequential regression trees. American journal of epidemiology. 2010;172(9):1070–6.

42. Doove LL, Van Buuren S, Dusseldorp E. Recursive partitioning for missing data imputation in the presence of interaction effects. Computational statistics & data analysis. 2014;72:92–104.

43. Shah DA, De Wolf ED, Paul PA, Madden LV. Predicting Fusarium head blight epidemics with boosted regression trees. Phytopathology. 2014 Jul;104(7):702–14.

44. Marshall A, Altman DG, Holder RL, Royston P. Combining estimates of interest in prognostic modelling studies after multiple imputation: current practice and guidelines. BMC Med Res Methodol. 2009 Dec;9(1):57.

45. Carugno M, Consonni D, Randi G, Catelan D, Grisotto L, Bertazzi PA, et al. Air pollution exposure, cause-specific deaths and hospitalizations in a highly polluted Italian region. Environmental Research. 2016 May;147:415–24.

46. Samoli E, Stafoggia M, Rodopoulou S, Ostro B, Declercq C, Alessandrini E, et al. Associations between fine and coarse particles and mortality in Mediterranean cities: results from the MED-PARTICLES project. Environmental health perspectives. 2013;121(8):932–8.

47. Scarinzi C, Chiusolo M, Alessandrini ER, Stafoggia M, Faustini A, Forastiere F, et al. Short-term effects of PM2. 5 and NO2 on mortality in 25 Italian cities: the EpiAir2 project. In 2013. p. 4996.

48. O’brien E, Masselot P, Sera F, Roye D, Breitner S, Ng CFS, et al. Short-term association between sulfur dioxide and mortality: a multicountry analysis in 399 cities. Environmental health perspectives. 2023;131(3):037002.

49. Martuzzi M, Mitis F, Iavarone I, Serinelli M. Health impact of PM10 and ozone in 13 Italian cities. WHO Regional Office for Europe. 2006;133.

50. Pope Iii CA, Burnett RT, Thun MJ, Calle EE, Krewski D, Ito K, et al. Lung cancer, cardiopulmonary mortality, and long-term exposure to fine particulate air pollution. Jama. 2002;287(9):1132–41.

51. Renzi M, Tinarelli G, Bauleo L, Maio S, Gariazzo C, Stafoggia M, et al. Short-term effects of PM10 on cause-specific mortality and the role of long-term environmental pressures in the industrial areas of Brindisi and Civitavecchia. Epidemiologia e Prevenzione. 2023;47(6):27–34.

52. Renzi M, Forastiere F, Calzolari R, Cernigliaro A, Madonia G, Michelozzi P, et al. Short-term effects of desert and non-desert PM10 on mortality in Sicily, Italy. Environment international. 2018;120:472–9.

53. Samoli E, Analitis A, Touloumi G, Schwartz J, Anderson HR, Sunyer J, et al. Estimating the exposure–response relationships between particulate matter and mortality within the APHEA multicity project. Environmental health perspectives. 2005;113(1):88–95.

54. Zauli Sajani S, Hänninen O, Marchesi S, Lauriola P. Comparison of different exposure settings in a case–crossover study on air pollution and daily mortality: counterintuitive results. Journal of exposure science & environmental epidemiology. 2011;21(4):385–94.

55. Faustini A, Alessandrini ER, Pey J, Perez N, Samoli E, Querol X, et al. Short-term effects of particulate matter on mortality during forest fires in Southern Europe: results of the MED-PARTICLES Project. Occupational and environmental medicine. 2015;72(5):323–9.

56. Chiusolo M, Cadum E, Stafoggia M, Galassi C, Berti G, Faustini A, et al. Short-term effects of nitrogen dioxide on mortality and susceptibility factors in 10 Italian cities: the EpiAir study. Environmental health perspectives. 2011;119(9):1233–8.

57. Kreyling W. Diverging long-term trends in ambient urban particle mass and number concentrations associated with emission changes caused by the German unification. Atmospheric Environment. 2003 Sep;37(27):3841–8.

58. Breitner S, Stölzel M, Cyrys J, Pitz M, Wölke G, Kreyling W, et al. Short-Term Mortality Rates during a Decade of Improved Air Quality in Erfurt, Germany. Environ Health Perspect. 2009 Mar;117(3):448–54.

59. Atkinson RW, Analitis A, Samoli E, Fuller GW, Green DC, Mudway IS, et al. Short-term exposure to traffic-related air pollution and daily mortality in London, UK. J Expo Sci Environ Epidemiol. 2016 Mar;26(2):125–32.

60. Cakmak S, Dales R, Kauri LM, Mahmud M, Van Ryswyk K, Vanos J, et al. Metal composition of fine particulate air pollution and acute changes in cardiorespiratory physiology. Environmental Pollution. 2014 Jun;189:208–14.

61. Eeftens M, Hoek G, Gruzieva O, Mölter A, Agius R, Beelen R, et al. Elemental Composition of Particulate Matter and the Association with Lung Function: Epidemiology. 2014 Sep;25(5):648–57.

62. Morishita M, Bard RL, Kaciroti N, Fitzner CA, Dvonch T, Harkema JR, et al. Exploration of the composition and sources of urban fine particulate matter associated with same-day cardiovascular health effects in Dearborn, Michigan. J Expo Sci Environ Epidemiol. 2015 Mar;25(2):145–52.

63. Peng RD, Bell ML, Geyh AS, McDermott A, Zeger SL, Samet JM, et al. Emergency Admissions for Cardiovascular and Respiratory Diseases and the Chemical Composition of Fine Particle Air Pollution. Environ Health Perspect. 2009 Jun;117(6):957–63.

64. Ibald-Mulli A, Wichmann HE, Kreyling W, Peters A. Epidemiological Evidence on Health Effects of Ultrafine Particles. Journal of Aerosol Medicine. 2002 Jun;15(2):189–201.

65. Kioumourtzoglou MA, Austin E, Koutrakis P, Dominici F, Schwartz J, Zanobetti A. PM2.5 and Survival Among Older Adults: Effect Modification by Particulate Composition. Epidemiology. 2015 May;26(3):321–7.

66. Penttinen P, Timonen KL, Tiittanen P, Mirme A, Ruuskanen J, Pekkanen J. Ultrafine particles in urban air and respiratory health among adult asthmatics. Eur Respir J. 2001 Mar;17(3):428–35.

67. Stölzel M, Breitner S, Cyrys J, Pitz M, Wölke G, Kreyling W, et al. Daily mortality and particulate matter in different size classes in Erfurt, Germany. J Expo Sci Environ Epidemiol. 2007 Aug 1;17(5):458–67.

68. Wichmann HE, Spix C, Tuch T, Wölke G, Peters A, Heinrich J, et al. Daily mortality and fine and ultrafine particles in Erfurt, Germany part I: role of particle number and particle mass. Res Rep Health Eff Inst. 2000 Nov;(98):5–86; discussion 87-94.

69. Seaton A. Hypothesis: Ill health associated with low concentrations of nitrogen dioxide--an effect of ultrafine particles? Thorax. 2003 Dec 1;58(12):1012–5.

70. Liu P, Ye C, Xue C, Zhang C, Mu Y, Sun X. Formation mechanisms of atmospheric nitrate and sulfate during the winter haze pollution periods in Beijing: gas-phase, heterogeneous and aqueous-phase chemistry. Atmos Chem Phys. 2020 Apr 7;20(7):4153–65.

71. Beelen R, Hoek G, Vienneau D, Eeftens M, Dimakopoulou K, Pedeli X, et al. Development of NO2 and NOx land use regression models for estimating air pollution exposure in 36 study areas in Europe – The ESCAPE project. Atmospheric Environment. 2013 Jun;72:10–23.

72. Di Q, Amini H, Shi L, Kloog I, Silvern R, Kelly J, et al. An ensemble-based model of PM2.5 concentration across the contiguous United States with high spatiotemporal resolution. Environment International. 2019 Sep;130:104909.

73. Van Donkelaar A, Martin RV, Brauer M, Boys BL. Use of Satellite Observations for Long-Term Exposure Assessment of Global Concentrations of Fine Particulate Matter. Environ Health Perspect. 2015 Feb;123(2):135–43.

