## Supplementary for "Disentangling non-linear and time-varying effects in assessing the short-term impact of air pollution on mortality: evidence from a 12-year study in a high-risk Italian area"

**Supplementary material**

**Environmental data preprocessing**

The air pollutant data for the period 2008-2019 were downloaded free of charge from the portal of the Regional Agency for Environmental Protection of the Tuscany region (ARPAT) (<https://www.arpat.toscana.it/datiemappe/dati/qualita-dellaria-dati-orari>). All available data underwent the entire validation cycle (<https://www.arpat.toscana.it/temi-ambientali/aria/monitoraggio/validazione-dati/ar_validazione_dati_bollettino.html>), but the monitoring network structure, station locations, and types of pollutants detected may vary over the years. During the period of interest, i.e., 2008 - 2019, monitoring stations were present in all the provinces of the Tuscany region: Arezzo (AR), Florence (FI), Grosseto (GR), Livorno (LI), Lucca (LU), Massa-Carrara (MS), Pisa (PI), Prato (PO), Pistoia (PT), Siena (SI), albeit with a non-uniform distribution across the territory, as depicted in Supplementary Figure S1a.

The temperature and humidity data, on the other hand, were obtained free of charge from the website of the Regional Hydrological Service (SIR, Servizio Idrologico Regionale) (<http://www.sir.toscana.it/consistenza-rete>) (Supplementary Fig. S1b). The humidity data collected by SIR were supplemented with those recorded by the LaMMA Consortium's monitoring stations (<https://www.lamma.toscana.it>) (Supplementary Fig. S1c).

Air pollutant daily time series

The monitoring stations across the Tuscany region can be equipped with sensors to detect the following air pollutants (not every station has all the sensors): Benzene (BENZENE), Methane (CH4), Carbon Monoxide (CO), Ethylbenzene (EBENZENE), Hydrogen Sulfide (H2S), Hydrogen Chloride (HCL), Meta-Xylene (M-XYLENE), Para-Xylene (MP-XYLENE), N-Heptane (N-EPTANO), N-Hexane (N-ESANO), N-Octane (N-OTTANO), Ammonia (NH3), Non-Methane Hydrocarbons (NMHC), Nitric Oxide (NO), Nitrogen Dioxide (NO2), Nitrogen Oxides (NOX), Ortho-Xylene (O-XYLENE), Ozone (O3), Particulate Matter with diameter less than 10 micrometers (PM10), Particulate Matter with diameter less than 2.5 micrometers (PM2.5), Sulfur Dioxide (SO2), Total Nitrogen (TN), Total Nitrogen Oxides (TNX), and Toluene (TOLUENE). Given the long observation period considered, i.e., (2008-2019), the monitoring stations have experienced periods of activity and inactivity, and some have been relocated from one area to another or undergone name changes. The data from 133 monitoring stations that had their names changed but did not undergo relocation were combined, and the suffix "_merged" was added to the name of the original monitoring station.

The frequency of sensor detection varies, and we executed several steps, depending on the pollutant, to obtain daily data (Supplementary Table S1). Specifically, for PM2.5 and PM10, we already had daily measurements, while for the other pollutants, we had hourly measurements. For all pollutants except CO and O3, the daily data was estimated as the average of the hourly measurements over 24 hours (if the percentage of missing observations within 24 hours exceeded 25%, the daily data was kept missing). For CO and O3, however, we first used a simple moving average procedure using equation (1):

$${sma}_{h}=\frac{1}{k}\sum_{i=0}^{k-1} x_{h+1}$$

(1)

where $x=x_{1},x_{2},\ldots, x_{h}, \ldots, x_{24}$ represents the hourly measurements of a day, $k$ is the width of the interval of interest (i.e., 8 hours), and $h=1, 2, \ldots, 17$ indicates the starting hour of the interval.

Similar to other pollutants, if the percentage of missing observations within the interval exceeded 25%, the moving average was set as missing. The values of the 17 moving averages within the day were then averaged to obtain the daily average (if the percentage of missing moving averages within 24 hours exceeded 25%, the daily data was kept missing).

Temperature and relative humidity daily time series

SIR acquired temperature data every 15 minutes. To derive the daily mean temperature, we followed the following procedure (Supplementary Figure S2): 1) we calculated the hourly average as the mean of the 4 measurements taken within the hour (only if all observations were missing, the hourly average was considered missing); 2) we calculated the tri-hourly average over disjoint and consecutive three-hour intervals (if more than one hourly observation was missing, the average was considered missing). The definition of these tri-hourly average values allowed us to define the following 8 parts of the day: night ([00 - 03], [03 - 06]), morning ([06 - 09], [09 - 12]), afternoon ([12 - 15], [15 - 18]), and evening ([18 - 21], [21 - 24]). The last step involves calculating the daily temperature as the mean of the 8 tri-hourly values (if the percentage of missing tri-hourly averages exceeded 25%, the daily data was kept missing). Calculating the daily average from tri-hourly averages rather than hourly ones helped us avoid underestimating the daily mean temperature (in case of missing morning and afternoon measurements) or overestimating it (in case of missing evening and night measurements). In fact, even in the absence of two consecutive tri-hourly values, e.g., morning ([06 - 09], [09 - 12]), the presence of afternoon measurements mitigated the underestimation effect of evening and night measurements. Conversely, in the absence of three or more tri-hourly observations (percentage of missing < 25%), the daily data was set to missing.

Regarding relative humidity data, daily minimum, mean, and maximum relative humidity values were freely downloaded from the SIR website. Since there were no measurements prior to the year 2020, we supplemented this information with relative humidity values measured by the LaMMA consortium's stations, which are less dense than the SIR network but provide data prior to 2020. To estimate the daily mean relative humidity, we followed the same procedure described earlier for raw temperature acquisitions.

In total, we reconstructed the daily time series of temperature and relative humidity data for 561 monitoring stations equipped with thermometers and hygrometers from the SIR network and 28 monitoring stations from the LaMMA consortium for humidity measurement.

**Pooling effect estimates and standard errors**

The pooling of effect estimates and standard errors was performed using Rubin’s multivariate rule. Specifically, the pooled estimate of the effect of PM10 (or PM2.5) ($\hat{\theta}_{pooled}$) was calculated as the arithmetic mean of the effect estimates obtained from each imputed dataset ($\hat{\theta}_{i}$ where $i$ ranges from 1 to 5), as follows: $\hat{\theta}_{pooled}=\frac{1}{5}\sum_{i=1}^{5} \hat{\theta}_{i}$. The pooled standard error is derived from components that account for both within- and between-imputation variance, reflecting the variability and uncertainty introduced by the multiple imputations. The within-imputation variance ($V_{W}$) is the mean of the variances within each imputed dataset, calculated as the average of the squared standard errors across the datasets: $V_{W}= \frac{1}{5}\sum_{i=1}^{5} \hat{SE}_{i}^{2}$, where $\hat{SE}_{i}^{2}$ represents the sampling variance of the parameter estimate in the $i^{th}$ imputed dataset. This measures the precision of the parameter of interest within each completed dataset. The between-imputation variance ($V_{B}$) quantifies the additional uncertainty arising from the missing data. It is calculated as the variance of the parameter estimates across the imputed datasets: $V_{B}=\frac{1}{5-1}\sum_{i=1}^{5} {(\hat{\theta}_{i}-\hat{\theta}_{pooled})}^{2}$. Finally, the total pooled standard error combines these components as follows: $\hat{SE}_{pooled}=\sqrt{V_{W}+V_{B}+ \frac{1}{5}V_{B}}$.

**Supplementary Table S1. Summary of detection frequencies and aggregation methods used to derive daily concentrations from hourly measurements for air pollutants.**

| **Air pollutant** | **Frequency of detection** | **Hourly to daily transformation** |
| --- | --- | --- |
| BENZENE | Hourly | Average |
| CH4 | Hourly | Average |
| CO | Hourly | SMA* and average |
| EBENZENE | Hourly | Average |
| H2S | Hourly | Average |
| HCL | Hourly | Average |
| M-XYLENE | Hourly | Average |
| MP-XYLENE | Hourly | Average |
| N-EPTANO | Hourly | Average |
| N-ESANO | Hourly | Average |
| N-OTTANO | Hourly | Average |
| NH3 | Hourly | Average |
| NMHC | Hourly | Average |
| NO | Hourly | Average |
| NO2 | Hourly | Average |
| NOX | Hourly | Average |
| O-XYLENE | Hourly | Average |
| O3 | Hourly | SMA and *average |
| PM10 | Daily | Average |
| PM2.5 | Daily | Average |
| SO2 | Hourly | Average |
| TN | Hourly | Average |
| TNX | Hourly | Average |
| TOLUENE | Hourly | Average |

SMA: simple moving average.

**Supplementary Table S2. Monitoring stations by pollutant, type, and geographic coordinates**

|  | **Monitoring stations** | | |
| --- | --- | --- | --- |
| **Air pollutant** | **Name** | **Type** | **Spatial coordinates (latitude, longitude)** |
| PM10 | MOSSE^¥^ | urban - traffic | 43.784807, 11.230453 |
|  | GRAMSCI | urban - traffic | 43.772067, 11.271167 |
|  | BASSI | urban - background | 43.785651, 11.286632 |
|  | BOBOLI | urban - background | 43.764213, 11.248074 |
|  | SIGNA* | urban - background | 43.793234, 11.097849 |
|  | SCANDICCI | urban - background | 43.755968, 11.191893 |
| PM2.5 | GRAMSCI° | urban - traffic | 43.772067, 11.271167 |
|  | BASSI° | urban - background | 43.785651, 11.286632 |
| NO2 | MOSSE^¥^ | urban - traffic | 43.784807, 11.230453 |
|  | GRAMSCI | urban - traffic | 43.772067, 11.271167 |
|  | SIGNA* | urban - background | 43.793234, 11.097849 |
|  | SCANDICCI | urban - background | 43.755968, 11.191893 |
| SO2 | MOSSE^ | urban - traffic | 43.784807, 11.230453 |
|  | BASSI | urban - background | 43.785651, 11.286632 |
|  | BOBOLI^ | urban - background | 43.764213, 11.248074 |

Based on Legislative Decree 155/2010, monitoring stations are classified according to the type of area in which they are located (urban, suburban, rural) and the station type, considering the dominant emission source (traffic, background, industrial).

*The SIGNA monitoring station was inactive during the 2011–2014 period.

^The SO2 sensors at the BOBOLI and MOSSE monitoring stations collected data only until 2012.

°PM2.5 was measured only from 2010 onward within the study area.

^¥^ The PM10 and NO2 sensors at the MOSSE monitoring station was inactive during the 2013.

**Supplementary Table S3. Number of days exceeding the 24-hour average threshold according to the 2021 WHO** guidelines (1). We **reported the average values over the multiple imputed datasets.**

| **Total number of days exceeding the 24-hour average threshold according to the 2021 WHO guidelines over the entire 12-year period** | | | | |
| --- | --- | --- | --- | --- |
| PM10 | PM2.5 | NO2 | SO2 |  |
| 441 | 1420.2 | 3848 | 0 |  |
| **Annual average number of days exceeding the 24-hour average threshold according to the 2021 WHO guidelines** | | | | |
| PM10 | PM2.5 | NO2 | SO2 |  |
| 36.75 | 142.02 | 320.666658 | 0 |  |
| **Number of days exceeding the 24-hour average threshold according to the 2021 WHO guidelines, year by year** | | | | |
| Year | PM10 | PM2.5 | NO2 | SO2 |
| 2008 | 81.6 |  | 363 | 0 |
| 2009 | 72.4 |  | 364.2 | 0 |
| 2010 | 55.8 | 160.4 | 364.8 | 0 |
| 2011 | 48 | 199.2 | 363.6 | 0 |
| 2012 | 43.2 | 176 | 358 | 0 |
| 2013 | 31 | 135.2 | 350 | 0 |
| 2014 | 17 | 106.2 | 301.6 | 0 |
| 2015 | 22.6 | 182 | 299.8 | 0 |
| 2016 | 21.4 | 117.8 | 286 | 0 |
| 2017 | 21 | 122.4 | 289.2 | 0 |
| 2018 | 14 | 114 | 261 | 0 |
| 2019 | 13 | 107 | 246.8 | 0 |

**Supplementary Table S4. Sensitivity analyses: percent change in natural, cardiovascular, and respiratory mortality associated with interquartile increases in air pollutant concentrations (with 90% confidence intervals), by lag structure. The interquartile range computed in the last year of observation (2019), specifically: 10.5** $\boldsymbol{\mu g}/{\boldsymbol{m}^{\boldsymbol{3}}}$ **for PM_10_, 7.5** $\boldsymbol{\mu g}/{\boldsymbol{m}^{\boldsymbol{3}}}$ **for PM_2.5_, 13.2** $\boldsymbol{\mu g}/{\boldsymbol{m}^{\boldsymbol{3}}}$ **for NO₂, and 0.98** $\boldsymbol{\mu g}/{\boldsymbol{m}^{\boldsymbol{3}}}$ **for SO₂.**

|  | **Natural deaths** | **Cardiovascular deaths** | **Respiratory deaths** |
| --- | --- | --- | --- |
| ***PM10*** | 0.46 (-0.20, 1.12) | -0.28 (-1.30, 0.75) | -0.16 (-2.12, 1.85) |
| Delayed (lag 2-5) | 0.62 (-0.12, 1.38) | 1.27 (0.05, 2.52) | 1.21 (-1.18, 3.65) |
| Prolonged (lag 0-5) | 0.85 (-0.01, 1.73) | 0.91 (-0.51, 2.35) | 0.93 (-1.83, 3.76) |
| ***PM2.5*** | 0.87 (0.17, 1.57) | 0.40 (-0.75, 1.57) | -0.58 (-2.78, 1.66) |
| Delayed (lag 2-5) | 0.85 (0, 1.71) | 1.55 ( 0.14, 2.98) | 0.82 (-1.85, 3.56) |
| Prolonged (lag 0-5) | 1.32 (0.33, 2.31) | 1.61 (-0.01, 3.24) | 0.32 (-2.73, 3.46) |
| ***NO2*** | 0.38 (-0.49, 1.25) | -0.34 (-1.79, 1.12) | -2.89 (-5.52, -0.2) |
| Delayed (lag 2-5) | 1.65 (0.54, 2.77) | 3.49 (1.63, 5.39) | -0.77 (-4.15, 2.74) |
| Prolonged (lag 0-5) | 1.67 (0.42, 2.94) | 2.74 (0.64, 4.90) | -2.63 (-6.37, 1.26) |
| ***SO2*** | 1.65 (0.92, 2.38) | 0.28 (-1.02, 1.59) | 3.69 (1.53, 5.90) |
| Delayed (lag 2-5) | 1.20 (0.45, 1.94) | 0.66 (-0.58, 1.93) | 2.73 (0.42, 5.08) |
| Prolonged (lag 0-5) | 1.70 (0.88, 2.53) | 0.63 (-0.76, 2.05) | 3.81 (1.31, 6.37) |

**Supplementary Table S5. Sensitivity analyses of age (< 65 years vs. ≥ 65 years) as an effect modifier of pollutant-related mortality (AIC values from models without interaction were always lower than those with interaction).**

|  |  | **Natural deaths** | **Cardiovascular deaths** | **Respiratory deaths** |
| --- | --- | --- | --- | --- |
| ***PM10*** | < 65 years | 2.81 (1.21, 4.44) | 2.88 (-0.64, 6.52) | 5.10 (-3.22, 14.15) |
|  | $\geq$ 65 years | 0.24 (-0.38, 0.87) | -0.43 (-1.43, 0.58) | -0.32 (-2.26, 1.66) |
| ***PM2.5*** | < 65 years | 1.63 (-0.26, 3.56) | 2.31 (-1.93, 6.73) | 0.11 (-9.81, 11.12) |
|  | $\geq$ 65 years | 0.80 (0.08, 1.52) | 0.32 (-0.85, 1.50) | -0.60 (-2.82, 1.66) |
| ***NO2*** | < 65 years | 4.21 (2.40, 6.05) | 3.31 (-0.63, 7.39) | 8.98 (-0.28, 19.11) |
|  | $\geq$ 65 years | 0.02 (-0.85, 0.91) | -0.51 (-1.97, 0.96) | -3.25 (-5.88, -0.55) |
| ***SO2*** | < 65 years | -0.62 (-2.24, 1.02) | 0.44 (-3.03, 4.03) | -5.54 (-13.68, 3.38) |
|  | $\geq$ 65 years | 1.85 (1.11, 2.59) | 0.27 (-1.05, 1.60) | 3.94 (1.77, 6.15) |

**Supplementary Table S6. Sensitivity analyses of different approaches to confounder modeling.**

|  | **Natural deaths** | **Cardiovascular deaths** | **Respiratory deaths** |
| --- | --- | --- | --- |
| ***PM10*** | 0.46 (-0.20, 1.12) | -0.28 (-1.30, 0.75) | -0.16 (-2.12, 1.85) |
| ns(calendar time, dof = 7/year) | 0.47 (-0.15, 1.09) | -0.14 (-1.15, 0.88) | -0.15 (-2.11, 1.84) |
| relative humidity_lag03 | 0.43 (-0.18, 1.04) | -0.27 (-1.26, 0.73) | -0.28 (-2.2, 1.68) |
| ns(relative humidity, df = 3) | 0.38 (-0.26, 1.02) | -0.24 (-1.28, 0.8) | -0.15 (-2.17, 1.9) |
| ns(relative humidity, df = 5) | 0.38 (-0.26, 1.02) | -0.24 (-1.28, 0.81) | -0.21 (-2.23, 1.84) |
| relative humidity + (relative humidity)^2^ | 0.38 (-0.26, 1.02) | -0.24 (-1.28, 0.81) | -0.15 (-2.17, 1.9) |
| relative humidity + (relative humidity)^2^ + relative humidity_lag1 + (relative humidity_lag1)^2^ + relative humidity_lag2 + (relative humidity_lag2)^2^ | 0.3 (-0.34, 0.94) | -0.33 (-1.38, 0.73) | -0.26 (-2.3, 1.82) |
| tps(relative humidity_lag1) | 0.44 (-0.17, 1.06) | -0.26 (-1.26, 0.75) | -0.36 (-2.29, 1.62) |
| ns(temperature_lag03, df = 6) | 0.44 (-0.17, 1.06) | -0.3 (-1.29, 0.7) | 0.03 (-1.91, 2.01) |
| ns(temperature, df = 5) | 0.62 (0.02, 1.23) | 0.15 (-0.83, 1.14) | -0.29 (-2.19, 1.65) |
| tps(temperature_lag1) | 0.48 (-0.13, 1.1) | -0.2 (-1.2, 0.8) | 0.03 (-1.9, 2) |
| ns(temperature, df = 6) + ns(temperature_lag3, df = 6) | 0.12 (-0.51, 0.76) | -0.48 (-1.51, 0.55) | -0.52 (-2.5, 1.5) |
| ***PM2.5*** | 0.87 (0.17, 1.57) | 0.40 (-0.75, 1.57) | -0.58 (-2.78, 1.66) |
| ns(calendar time, dof = 7/year) | 0.68 (-0.04, 1.4) | 0.3 (-0.88, 1.5) | -1.06 (-3.31, 1.23) |
| relative humidity_lag03 | 0.83 ( 0.13, 1.53) | 0.41 (-0.73, 1.57) | -0.77 (-2.95, 1.47) |
| ns(relative humidity, df = 3) | 0.77 (0.03, 1.5) | 0.66 (-0.55, 1.88) | -0.99 (-3.28, 1.36) |
| ns(relative humidity, df = 5) | 0.76 (0.02, 1.5) | 0.61 (-0.59, 1.84) | -1.04 (-3.33, 1.31) |
| relative humidity + (relative humidity)^2^ | 0.77 (0.04, 1.51) | 0.66 (-0.55, 1.88) | -0.99 (-3.28, 1.35) |
| relative humidity + (relative humidity)^2^ + relative humidity_lag1 + (relative humidity_lag1)^2^ + relative humidity_lag2 + (relative humidity_lag2)^2^ | 0.62 (-0.12, 1.37) | 0.53 (-0.69, 1.76) | -1.16 (-3.48, 1.22) |
| tps(relative humidity_lag1) | 0.84 (0.14, 1.55) | 0.42 (-0.74, 1.59) | -0.75 (-2.95, 1.51) |
| ns(temperature_lag03, df = 6) | 0.84 (0.13, 1.55) | 0.31 (-0.85, 1.49) | -0.22 (-2.45, 2.06) |
| ns(temperature, df = 5) | 1.13 (0.44, 1.82) | 0.98 (-0.15, 2.13) | -0.69 (-2.84, 1.52) |
| tps(temperature_lag1) | 0.93 (0.23, 1.65) | 0.52 (-0.64, 1.69) | -0.46 (-2.69, 1.81) |
| ns(temperature, df = 6) + ns(temperature_lag3, df = 6) | 0.63 (-0.09, 1.36) | 0.26 (-0.93, 1.47) | -0.58 (-2.87, 1.76) |
| ***NO2*** | 0.38 (-0.49, 1.25) | -0.34 (-1.79, 1.12) | -2.89 (-5.52, -0.2) |
| ns(calendar time, dof = 7/year) | 0.23 (-0.64, 1.11) | -0.45 (-1.91, 1.02) | -3.21 (-5.85, -0.51) |
| relative humidity_lag03 | 0.31 (-0.55, 1.19) | -0.31 (-1.75, 1.15) | -3.19 (-5.8, -0.51) |
| ns(relative humidity, df = 3) | 0.31 (-0.59, 1.22) | -0.22 (-1.72, 1.31) | -3.21 (-5.94, -0.41) |
| ns(relative humidity, df = 5) | 0.3 (-0.6, 1.21) | -0.22 (-1.73, 1.31) | -3.09 (-5.83, -0.28) |
| relative humidity + (relative humidity)^2^ | 0.3 (-0.6, 1.21) | -0.22 (-1.72, 1.31) | -3.23 (-5.96, -0.43) |
| relative humidity + (relative humidity)^2^ + relative humidity_lag1 + (relative humidity_lag1)^2^ + relative humidity_lag2 + (relative humidity_lag2)^2^ | 0.1 (-0.82, 1.03) | -0.48 (-2.02, 1.09) | -3.2 (-6.07, -0.4) |
| tps(relative humidity_lag1) | 0.34 (-0.53, 1.21) | -0.29 (-1.74, 1.17) | -3.18 (-5.8, -0.49) |
| ns(temperature_lag03, df = 6) | 0.34 (-0.53, 1.22) | -0.4 (-1.85, 1.08) | -2.52 (-5.18, 0.21) |
| ns(temperature, df = 5) | 0.42 (-0.45, 1.3) | 0.01 (-1.45, 1.49) | -3 (-5.64, -0.29) |
| tps(temperature_lag1) | 0.39 (-0.49, 1.27) | -0.3 (-1.75, 1.18) | -2.74 (-5.39, -0.03) |
| ns(temperature, df = 6) + ns(temperature_lag3, df = 6) | 0.03 (-0.87, 0.93) | -0.56 (-2.05, 0.96) | -3.06 (-5.75, -0.3) |
| ***SO2*** | 1.65 (0.92, 2.38) | 0.28 (-1.02, 1.59) | 3.69 (1.53, 5.90) |
| ns(calendar time, dof = 7/year) | 1.69 (0.91, 2.48) | 0.07 (-1.34, 1.50) | 3.78 (1.40, 6.22) |
| relative humidity_lag03 | 1.58 (0.86, 2.31) | 0.3 (-0.99, 1.6) | 3.36 (1.22, 5.54) |
| ns(relative humidity, df = 3) | 1.63 (0.9, 2.36) | 0.35 (-0.95, 1.67) | 3.53 (1.37, 5.74) |
| ns(relative humidity, df = 5) | 1.64 (0.91, 2.37) | 0.35 (-0.96, 1.68) | 3.47 (1.31, 5.67) |
| relative humidity + (relative humidity)^2^ | 1.63 (0.9, 2.36) | 0.35 (-0.95, 1.67) | 3.53 (1.37, 5.73) |
| relative humidity + (relative humidity)^2^ + relative humidity_lag1 + (relative humidity_lag1)^2^ + relative humidity_lag2 + (relative humidity_lag2)^2^ | 1.6 (0.87, 2.33) | 0.27 (-1.04, 1.59) | 3.68 (1.51, 5.89) |
| tps(relative humidity_lag1) | 1.61 (0.88, 2.34) | 0.31 (-0.98, 1.62) | 3.43 (1.29, 5.62) |
| ns(temperature_lag03, df = 6) | 1.65 (0.92, 2.39) | 0.29 (-1.01, 1.61) | 3.75 (1.58, 5.97) |
| ns(temperature, df = 5) | 1.79 (1.06, 2.53) | 0.58 (-0.72, 1.9) | 3.48 (1.33, 5.67) |
| tps(temperature_lag1) | 1.74 (1, 2.48) | 0.43 (-0.86, 1.74) | 3.72 (1.55, 5.93) |
| ns(temperature, df = 6) + ns(temperature_lag3, df = 6) | 1.59 (0.86, 2.32) | 0.32 (-0.98, 1.64) | 3.51 (1.35, 5.72) |

**Supplementary Table S7. Sensitivity analyses: linear effect of average pollutant values measured only by monitoring stations inactive for less than one consecutive year.**

|  | **Natural deaths** | **Cardiovascular deaths** | **Respiratory deaths** |
| --- | --- | --- | --- |
| ***PM10*** | 0.62 (-0.05, 1.3) | 0.04 (-1.06, 1.15) | -0.2 (-2.32, 1.96) |
| ***NO2*** | 0.22 (-0.61, 1.06) | -0.24 (-1.62, 1.17) | -3.2 (-5.76, -0.57) |
| ***SO2*** | 1.42 (0.79, 2.06) | 0.34 (-0.75, 1.45) | 2.76 (0.82, 4.74) |

| 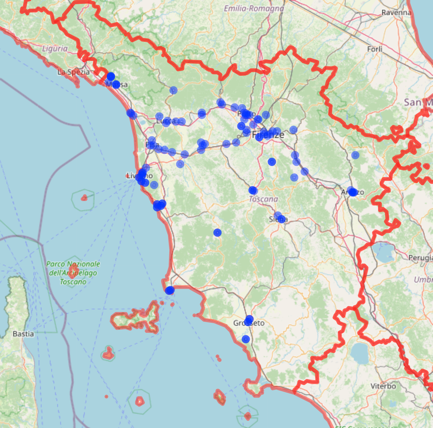  **a** | 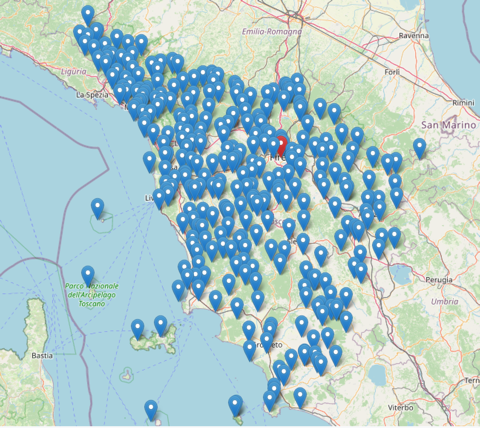  **b** | 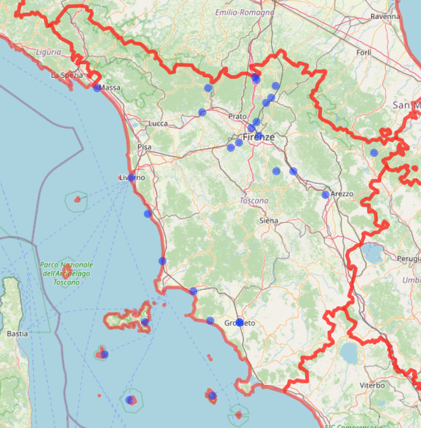  **c** |
| --- | --- | --- |

**Supplementary Figure S1. Spatial distribution of monitoring stations in Tuscany in 2019: air pollutants monitoring station (a), SIR monitoring stations (b), and LaMMA monitoring stations (c).**

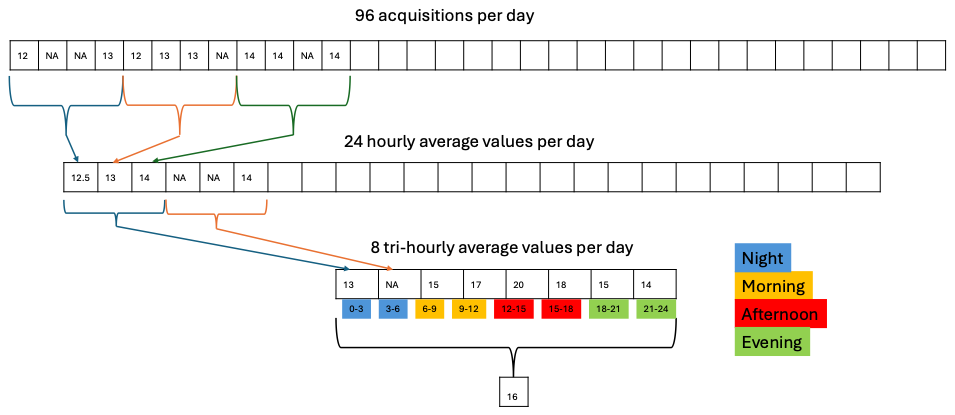
**Supplementary figure S2. Temperature data processing: from raw measurements to daily average values.**

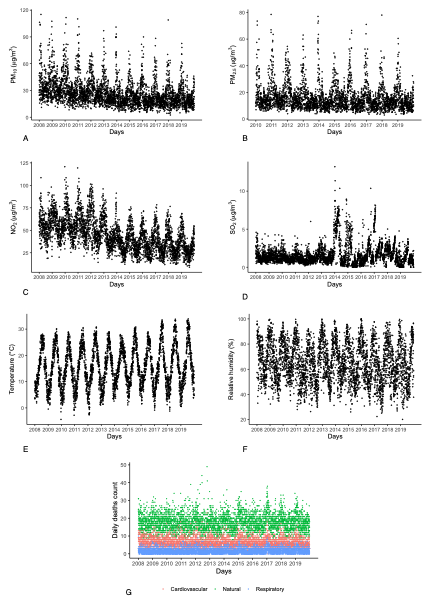

**Supplementary figure S3.** Historical time series for daily concentration levels for PM_10_ (A), PM_2.5_ (B), NO_2_ (C), SO_2_ (D), mean temperature (E), mean relative humidity (F) and mortality (G).

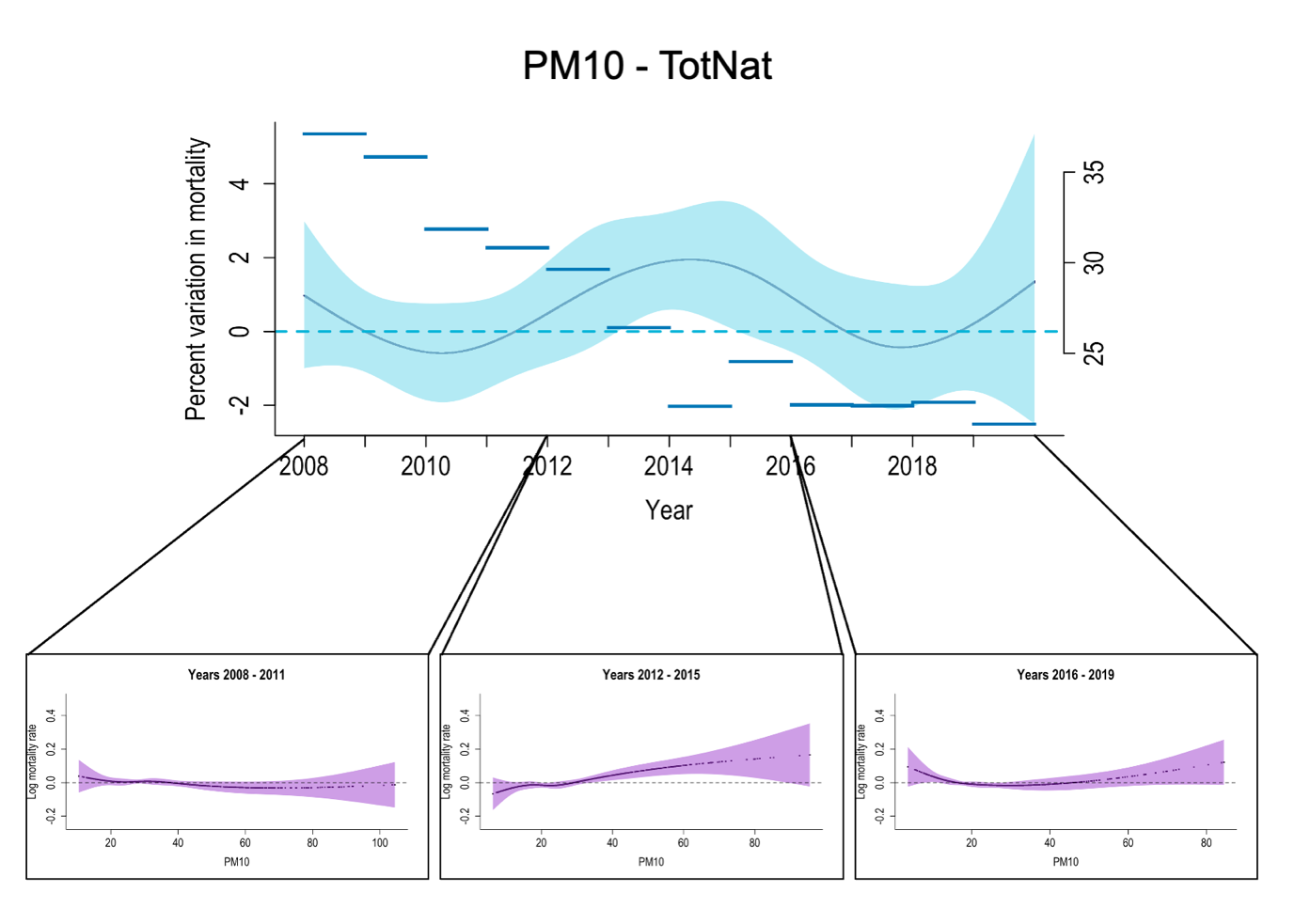

**Supplementary figure S4.** Time-variant linear effects of PM_10_on natural mortality. For each four-year period, the non-linear dose-response curve estimated over the corresponding period is reported.

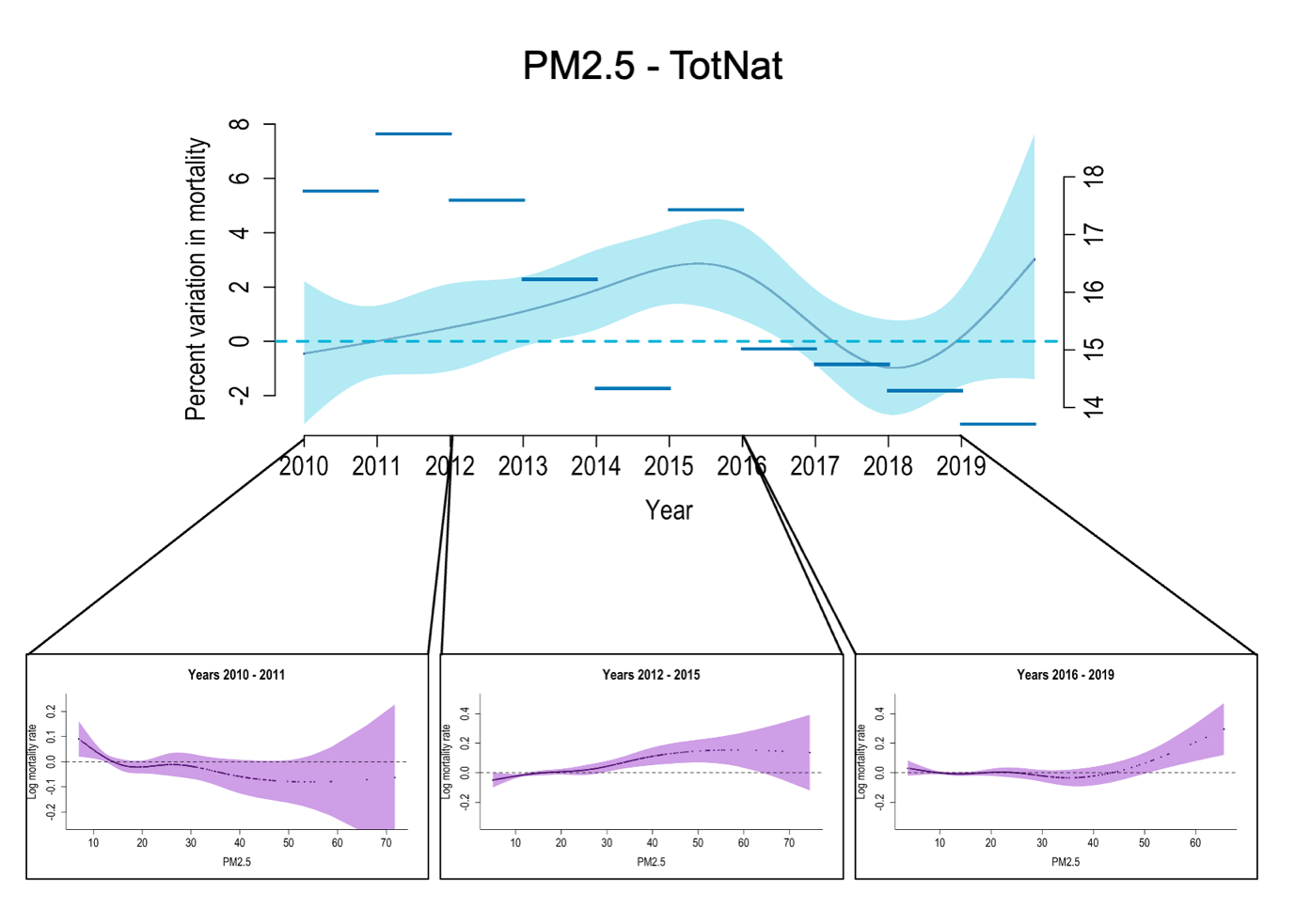

**Supplementary figure S5.** Time-variant linear effects of PM_2.5_ on natural mortality. For each four-year period, the non-linear dose-response curve estimated over the corresponding period is reported.

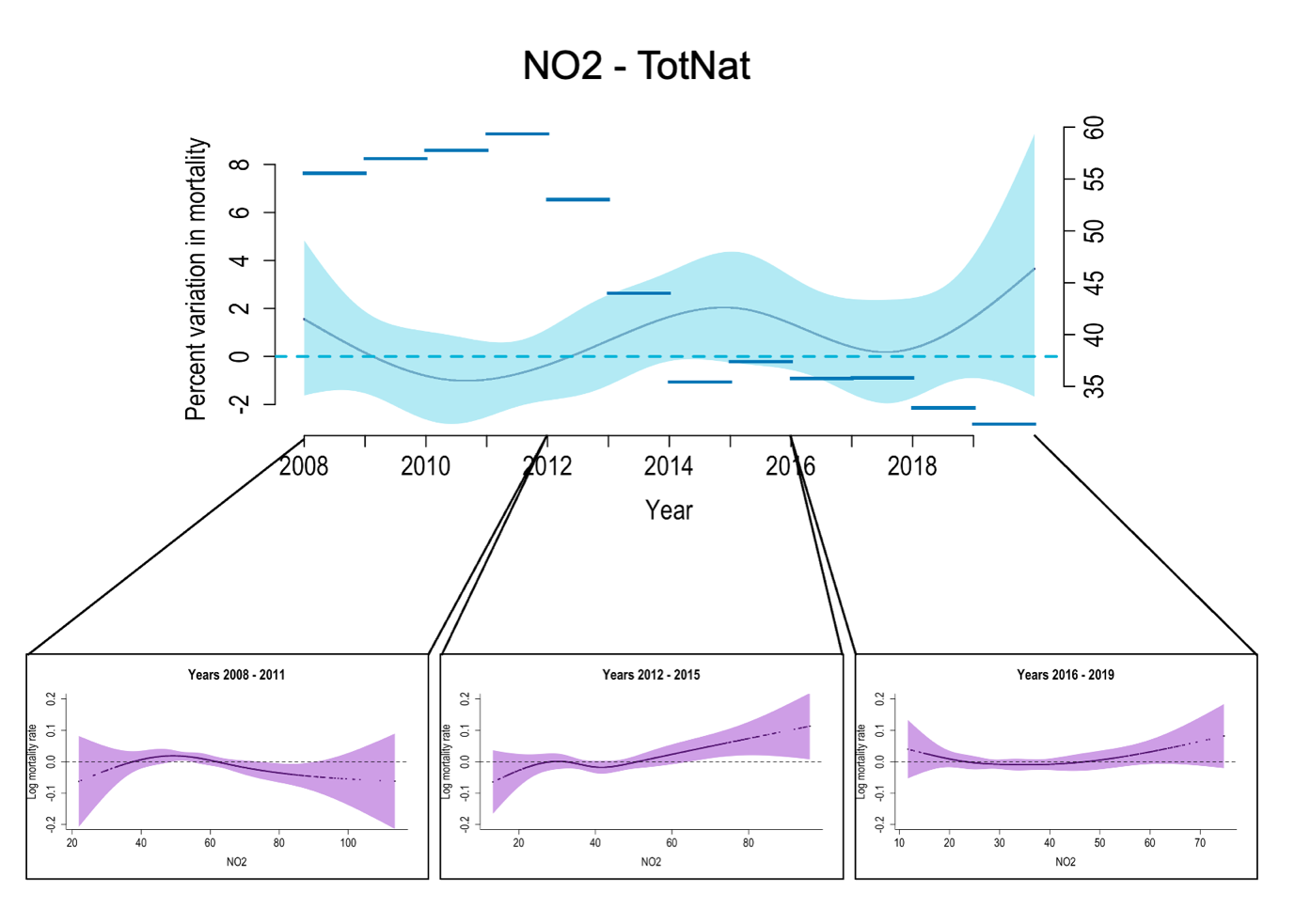

**Supplementary figure S6.** Time-variant linear effects of NO_2_ on natural mortality. For each four-year period, the non-linear dose-response curve estimated over the corresponding period is reported.

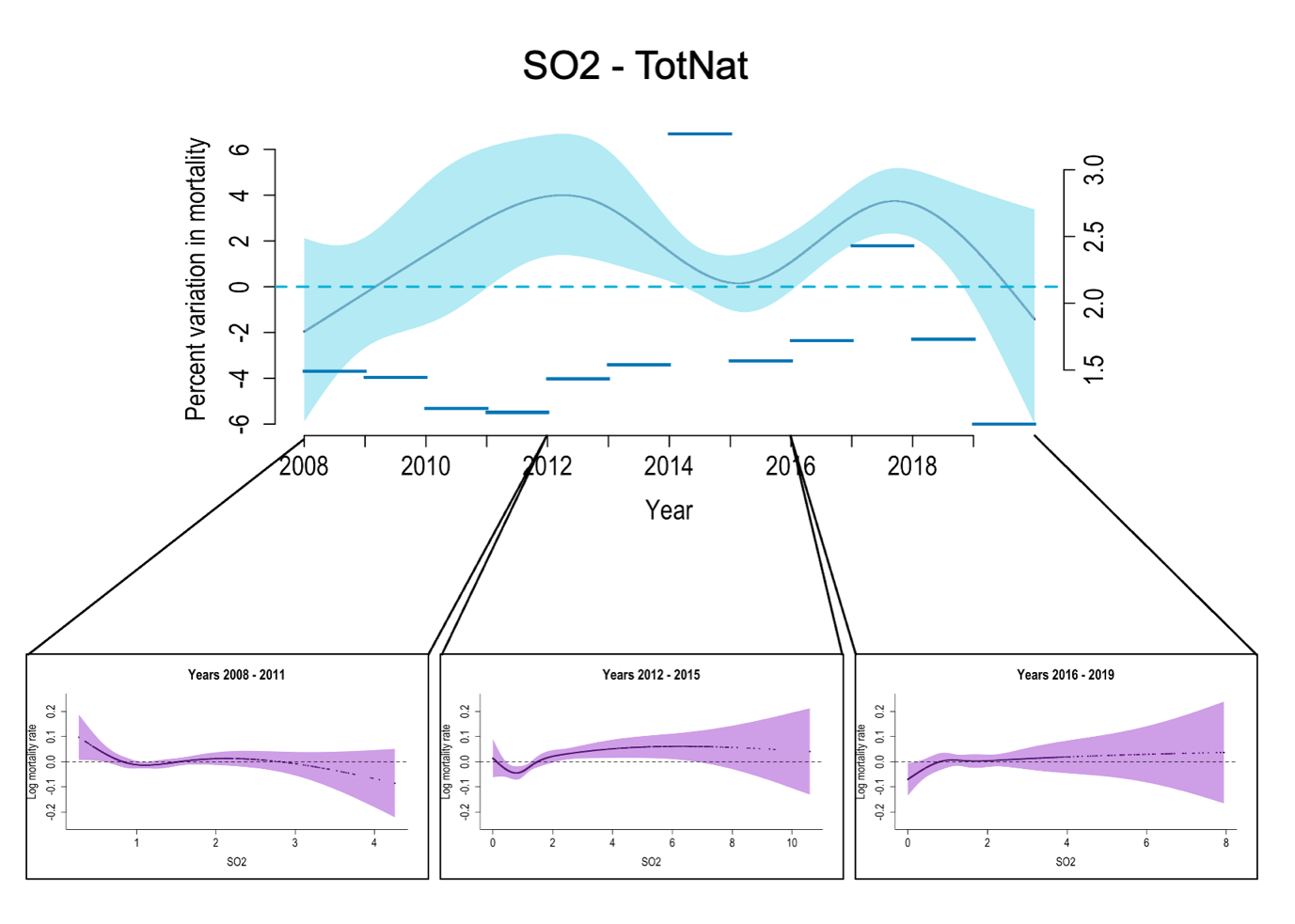

**Supplementary figure S7.** Time-variant linear effects of SO_2_ on natural mortality. For each four-year period, the non-linear dose-response curve estimated over the corresponding period is reported.

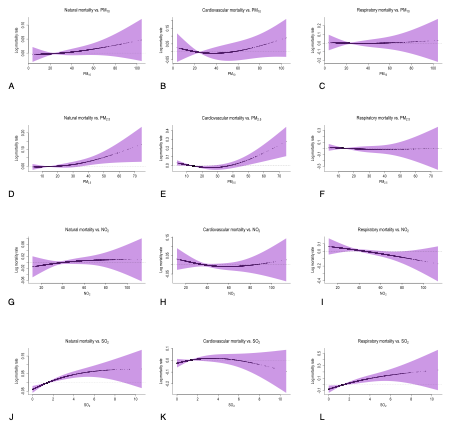
**Supplementary figure S8. Non-linear effects of PM10, PM2.5, NO2, and SO2 on mortality, using three internal knots positioned at the quartiles of the pollutant distribution.**

**References**

1.	WHO Global Air Quality Guidelines: Particulate Matter (PM2. 5 and PM10), Ozone, Nitrogen Dioxide, Sulfur Dioxide and Carbon Monoxide. 1st ed. Geneva: World Health Organization; 2021. 1 p.
